# Quantifying and identifying strategies to improve rotavirus vaccine impact in low- and middle-income countries

**DOI:** 10.64898/2026.05.12.26353059

**Authors:** Ernest O. Asare, Jiye Kwon, Xiao Li, Mohammad A. Al-Mamun, Belinda L. Lartey, Khuzwayo C. Jere, Nigel A. Cunliffe, George E. Armah, Benjamin A. Lopman, Virginia E. Pitzer

## Abstract

**Background:** Robust quantitative evidence on the impact of rotavirus vaccines, their potential benefits in countries without vaccination, and strategies to improve performance in low- and middle-income countries (LMICs) is essential for informing policy decisions aimed at sustaining and expanding vaccination programs.

**Methods and Findings:** We used an age-structured compartmental model of rotavirus gastroenteritis (RVGE) transmission that accounts for the natural history of infection to estimate vaccine impact across 112 LMICs. The model incorporates country-specific data on demographics, transmission dynamics, vaccination schedules, coverage levels, and vaccine performance. We simulated multiple scenarios, including the continuation of current vaccination programs, vaccine introduction in countries without programs, the addition of a third dose, scale-up of coverage to 95% in low-coverage settings, and suspension of vaccination. We quantified health impacts by estimating cases, deaths, and disability-adjusted life years (DALYs) averted from 2006 to 2024 and projected over 2025 to 2034 using either no vaccination or the current program as counterfactual. We estimated that rotavirus vaccination averted a median of 268 million RVGE cases (95% uncertainty interval [UI]: 228-306 million), 35 million moderate-to-severe cases (95% UI: 30-38 million), 817 thousand deaths (95% UI: 684-928 thousand), and 53 million DALYs (95% UI: 45-61 million) between 2006 and 2024, resulting from 81 countries with vaccination programs out of 112 LMICs. Using the current vaccination as a baseline, we estimated substantial additional benefits for all strategies, except for suspension, which would increase the RVGE burden over the next 10 years. Scaling up coverage to at least 95% across all 112 LMICs, with countries without the vaccine using the 6/10/14-week schedule, could avert a median of 296 million RVGE cases (95% UI: 243-358 million), 832 thousand deaths (95% UI: 694-932 thousand), and 55 million DALYs (95% UI: 45-61 million), respectively. Furthermore, adding a third dose in the 51 countries currently using a two-dose schedule could enhance vaccine impact, averting a median of 123 million RVGE cases (95% UI: 102-145 million), 377 thousand deaths (95% UI: 310-440 thousand), and 24 million DALYs (95% UI: 20-28 million), respectively, compared to the two-dose schedule.

**Conclusions:** Our model demonstrates that rotavirus vaccination provides substantial health benefits, with an even greater impact achievable through broader adoption and increased coverage. Adding a third dose to the standard two-dose Rotarix schedule could be an additional strategy to improve vaccine impact in LMICs. These findings support continued efforts to sustain and expand vaccination programs across LMICs. The country-specific, model-estimated rotavirus burden can also inform economic evaluations to guide more effective vaccination strategies.

## Introduction

Since 2009, the World Health Organization (WHO) has recommended rotavirus vaccines as the most effective strategy for reducing the burden of rotavirus gastroenteritis (RVGE) [1]. There are currently four licensed live-attenuated oral rotavirus vaccines prequalified by the WHO for global use (RotaTeq, Rotarix, Rotavac, and RotaSiil), and several next-generation rotavirus vaccines are in development [2, 3]. There has been an extensive global effort to expand rotavirus vaccination in many national immunization programs, with a total of 135 countries having implemented the rotavirus vaccine as of July 2025 [4]. This continuous global expansion of rotavirus vaccines has resulted in significant reductions in RVGE hospitalizations and mortality [5–9]. Furthermore, global RVGE morbidity and mortality are expected to decrease even further, as several countries accounting for a large proportion of the global rotavirus burden have recently introduced or are planning to introduce the vaccine [8, 9]. Despite the substantial impact of rotavirus vaccines, there are significant differences in vaccine impact between high-income countries and low- and middle-income countries (LMICs) [5,10,11].

Given the suboptimal performance of rotavirus vaccines in LMICs, understanding key drivers and identifying strategies to enhance effectiveness is critical for policymakers. Some proposed strategies include modifying the dosing schedule, adding a third dose to the standard two-dose regimen, and administering a booster dose at 9 months of age [3, 12–16]. With the removal of age restrictions for the rotavirus vaccine by the WHO [17], some of these strategies can more readily be incorporated into countries rotavirus immunization schedules. These strategies are difficult to evaluate using surveillance data because their effects can only be measured after implementation, limiting the ability to compare vaccination schedules before policy decisions are made. Mathematical modeling can therefore be employed to assess and quantify their impact at the country level.

Numerous dynamic models of rotavirus transmission have been extensively used to evaluate the country-specific and multi-country impacts of rotavirus vaccines [15, 16, 18–24]. These models provide quantitative evidence of reductions in RVGE-related hospitalizations and mortality in LMICs, despite the suboptimal performance of the vaccine. A major limitation of most of these modeling studies is that they primarily evaluate the impact of a single vaccination strategy (i.e., the implemented vaccine schedule), using no vaccination as the baseline [18, 20, 25]. Current mathematical modeling studies that have quantified the impact of different vaccine schedules or vaccine switching are limited to single countries [15,16,24]. Consequently, there is a lack of modeling framework that can be used to quantify the relative health impact of different vaccination strategies across the LMICs. These comparative vaccine impact studies are relevant to policy decisions regarding switching to alternative vaccine strategies or changing dosing schedules.

We aimed to quantify the overall and country-specific impact of the historical rotavirus vaccination program in 81 of the 112 LMICs that had introduced the vaccine by the end of 2024. Furthermore, we project the impact across all 112 LMICs from 2025 to 2034 under various vaccination strategies. Finally, we develop a modeling framework addressing key policy questions, including: What is the historical impact of rotavirus vaccination in 112 LMICs? What is the projected impact with increased coverage? What would be the total impact if all 112 LMICs introduced the vaccine at high coverage? What would be the impact of switching the vaccine dosing regimen? This analysis provides policymakers with robust health impact estimates and informs strategies to improve vaccine effectiveness across LMICs.

## Methods

### Rotavirus vaccination status and coverage

Across the 112 LMICs, 83 have introduced rotavirus vaccination as of July 2025 (Fig 1A). The first introduction occurred in 2006, and two additional countries adopted the vaccine in 2025. Currently, most countries (52) primarily use the two-dose Rotarix vaccine, with a comparable number of countries using the three-dose vaccines: RotaTeq (7 countries), Rotavac (13), and Rotasiil (9). India administers both Rotavac and Rotasiil vaccines, while Uzbekistan uses both RotaTeq and Rotarix. Vaccine coverage varies substantially across countries. From the year of each country’s introduction through 2024, the average coverage of the last dose ranged from 41% in Indonesia to 99% in Tonga (Fig 1B). The coverage remains suboptimal in many countries: 30% have coverage below 70%, only 20% exceed 90%, and overall rotavirus vaccine coverage averages approximately 70% [26]. For the 10-year projection period (2025-2034), we used each country’s 2024 rotavirus vaccine coverage or DTP3 coverage (for countries without rotavirus vaccination) as input (S1 Fig, S1 Appendix).

**Fig 1.**
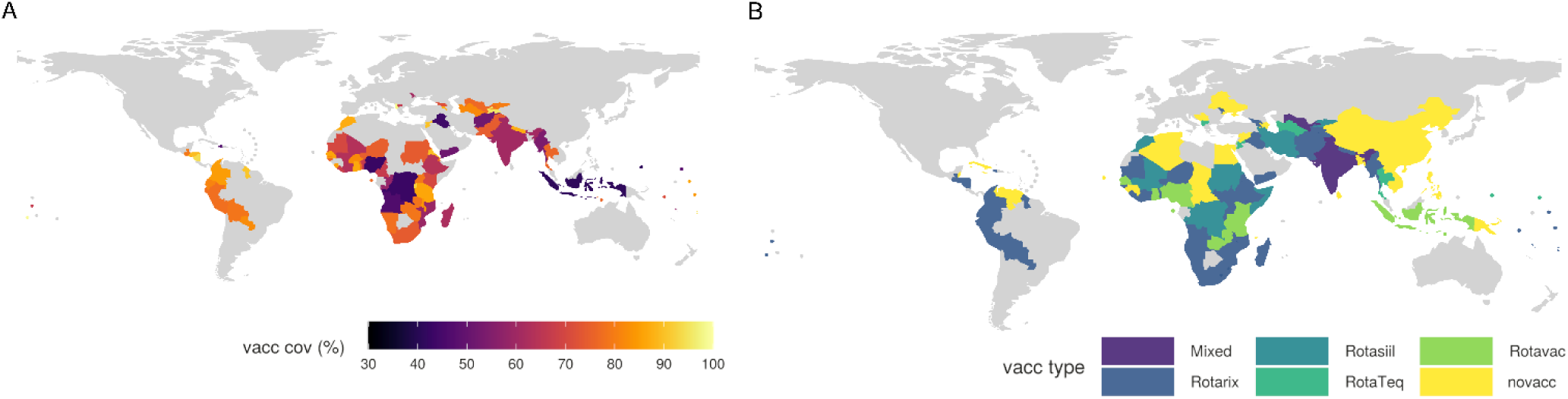
Country-specific rotavirus vaccination coverage and vaccine product type. **(A) Average rotavirus vaccination coverage from the year of introduction through 2024 for each country, used in retrospective simulations.** (B) The type of rotavirus vaccine (Rotarix, Rotavac, Rotasiil, and RotaTeq) used in each country with an existing vaccination program. Data sourced from the WHO/UNICEF Estimates of National Immunization Coverage (WUENIC) [26].

### Country-specific demographic input data

We used country-specific baseline population structures, crude annual birth and death rate data from the U.S. Census Bureau [27] to parameterize population dynamics. Average country-specific birth rates between 2006 and 2024 and the 2025 and 2034 projection period are shown in S2 Fig.

### Model description

We used our well-established and validated deterministic, age-stratified dynamic compartmental model of RVGE transmission [15,16,21–24,28,29]. The model realistically reflects the natural history of RVGE infection, vaccine response, and the duration of vaccine-induced immunity. A detailed model description, schematic diagram (S3 Fig), and fixed model parameters (S1 Table) are provided in S1 Text. The key assumptions underlying the model are: (i) newborns are born with maternally derived immunity, which wanes, rendering them susceptible to primary RVGE infection; (ii) individuals can be infected multiple times, with the severity of infection decreasing with each subsequent infection; (iii) following infection, individuals gain temporary immunity, which wanes before they become susceptible to reinfection; and (iv) infections that occur subsequent to the tertiary infection are typically asymptomatic or mild.

To account for vaccination, we introduced a vaccinated compartment, with the rate of movement from unvaccinated to vaccinated based on first-dose coverage. We made the following assumptions: (i) infants immunized with the vaccine gain temporary immunity to reinfection, similar to natural infection; (ii) after vaccine-induced immunity wanes following the first, second, and third doses, infants become susceptible to primary, secondary, and tertiary infections, respectively; (iii) those who failed to respond to the first dose have a lower probability of responding to subsequent doses; and (iv) the duration of vaccine-induced immunity is the same for all doses.

### Estimation of country-specific R_0_ values

To estimate country-specific values of the *R_0_* across the 112 LMICs, we used regression-based model estimates of the mean age of severe RVGE infection [18]. This regression model incorporates key variables known to influence rotavirus transmission dynamics, including gross domestic product, birth rate, under-five mortality rate, and the proportion of the population living in rural areas. The model has been validated and demonstrates strong predictive performance [18]. We used country-specific demographic data to simulate our RVGE transmission model in the absence of vaccination. We extracted the corresponding model-estimated ages of first infection across a range of *R_0_* values. We estimated the country-specific *R_0_* values and their 95% confidence intervals (CIs) by comparing the model-simulated age of first infection with that from the regression model. We subsequently generated 1,000 samples within the country-specific estimated *R_0_* 95% CIs, assuming a normal distribution for our simulations. The estimated country-specific *R_0_* and its 95% CIs are provided in S1 Appendix, and the spatial distribution of the mean is shown in S4 Fig.

### Estimation of country-specific vaccine response rates

To obtain country-specific vaccine response rates, we developed a simple linear regression model to assess the relationship between vaccine response rates and vaccine efficacy (S5 Fig). We utilized all published data on the efficacy and response rates of available rotavirus vaccines from clinical trials to conduct this analysis [11]. Using the identified relationship between vaccine response rates and efficacy, we estimated country-specific response rates for both two-dose and three-dose vaccines, based on the published country-specific predicted vaccine efficacy from Prunas et al. [11]. The probability of responding to each vaccine dose was estimated following the formulation of Pitzer et al. [15]. A detailed description of the approach and the resulting country-specific data used for the model is provided in the S1 Text and the S1 Appendix, respectively.

### Estimation of non-country-specific model parameters

In our model, we incorporated non-country-specific parameters (see S1 Appendix) assumed to be broadly applicable across settings. These included the duration of maternal immunity, the proportion of moderate-to-severe diarrhea cases hospitalized, the proportion of subsequent infections that are severe, and the duration of vaccine-induced immunity. Parameter estimates were derived from separate fits of the same model to pre- and post-vaccination surveillance data from three sites in Ghana, as well as from Malawi and Bangladesh [15, 16, 21–24]. For each parameter, we sampled 1,000 values from a normal distribution defined by the lowest and highest 95% CIs bounds across all fits to capture a wider uncertainty range. For vaccine-induced immunity, we used model-estimated values derived from fitting the same model separately to post-Rotarix vaccine introduction RVGE cases in Malawi and Ghana and post-Rotavac vaccine introduction in Ghana [15, 16, 22–24]. To account for potential variations in the duration of vaccine-induced immunity across different vaccines and settings, we conducted 1,000 simulations assuming a uniform distribution. These simulations were based on the lowest and highest 95% CIs from the model estimates. This approach provides robust and generalizable parameter estimates for the duration of vaccine-induced immunity.

### Historical impact of rotavirus vaccination program

To estimate the impact of historical rotavirus vaccination programs in countries that have implemented the vaccine, we compared model simulations including vaccination to a no-vaccination baseline. For each country, the model was run using country-specific vaccine response rates, dosing schedules, and annual vaccine coverage data. Outputs from the vaccination scenario were extracted starting from the year of vaccine introduction and compared to corresponding no-vaccination simulations over the same period.

### Projection scenarios over a 10-year period (2025–2034)

To explore and quantify the impact of rotavirus vaccination across 112 LMICs, we simulated the model under five vaccination scenarios over a 10-year period (2025-2034):

i. Current strategy scenario: We simulated the model across all 112 LMICs. Countries (83 as of 2025) with existing rotavirus vaccination programs maintained their current schedules and coverage levels, while no introduction was assumed for countries without vaccination programs.
ii. Full adoption scenario: Countries with existing programs maintained their current schedules and coverage. In the 29 countries without rotavirus vaccination, we assumed the introduction of either a three-dose schedule (administered at 6, 10, and 14 weeks) or a two-dose schedule (administered at 6 and 10 weeks or 10 and 14 weeks), using country-specific DTP3 coverage as a proxy for uptake.
iii. Scaled-up coverage to 95% scenario: We assessed the impact of increasing rotavirus vaccine coverage to 95% in countries where current rotavirus or DTP3 coverage remains below this threshold.
iv. Additional dose strategy scenario: This scenario assessed the impact of adding a third dose (administered at either 6 or 14 weeks) in the 51 countries currently using a two-dose schedule.
v. Suspension scenario: This is a worst-case scenario in which rotavirus vaccination programs are suspended in countries with existing programs, and no introductions occur in countries without vaccination. Given that some countries have already suspended their vaccination programs, and countries are facing financial constraints under Gavi 6.0, providing evidence of the adverse health impacts of program suspension is still relevant for public health decision-making.

We assessed the impact of historical and projected rotavirus vaccination by estimating the number of cases, deaths, and disability-adjusted life years (DALYs). Using no vaccination (for the retrospective period, 2006-2024) and the current vaccination program (projected period, 2025-2034), we quantify the various strategies as the percent change of averted cases, deaths, and DALYs.

### Estimation of deaths and disability-adjusted life years (DALYs)

The rotavirus-specific case fatality rate (CFR) was estimated using data from a published systematic review (Asare et al., [30]) and stratified by WHO region and age group (<1 year and 1–4 years). Details of the CFR estimation approach and values are provided in the S1 Text. We assumed a uniform CFR across all countries within each WHO region and generated 1,000 samples from a beta distribution to account for uncertainty. To estimate the number of deaths in each age group and country, the model-simulated number of cases was multiplied by the corresponding country-specific CFR stratified by WHO regions (see S2 Table). The estimated deaths and model-simulated cases were then used to calculate age-stratified years of life lost (YLLs) and years lived with disability (YLDs) for each country.

We estimated YLLs due to premature death using life expectancy at birth data from the U.S. Census Bureau [27]. For each country, YLLs were calculated by subtracting the average age at death (approximated as the mean age of moderate-to-severe RVGE cases) from the country-specific life expectancy at birth and multiplying the result by the number of model-simulated deaths in each age group.

The YLDs were estimated by multiplying the country-specific number of moderate-to-severe RVGE cases by the average duration of infection and the disability weight for diarrhea [31]. The duration of infection and the disability weight were assumed to be the same across both age groups (see S1 Text). To account for uncertainty, we generated 1,000 samples from a beta distribution defined by the disability weights for moderate and severe diarrhea (see S1 Text). Finally, the DALYs were then calculated as the sum of YLLs and YLDs across countries and WHO regions.

### Model validation

We conducted a retrospective model validation to assess the predictive performance of our model across LMICs. Country-specific model predictions in the absence of vaccination were validated using regression-based estimates of 2016 rotavirus cases in children under 5 years of age from the Global Burden of Disease study, serving as a proxy for observational data (see Troeger et al. 2018 [32]). Due to the lack of country-specific post-vaccination observational data across most LMICs, we were unable to validate post-vaccination predictions globally. However, the model has been extensively validated in Ghana and Malawi, where post-vaccination data are available. In these settings, the model has demonstrated strong predictive performance and produced realistic estimates under various conditions, including the impact of COVID-19 in Malawi and the switch from Rotarix to Rotavac in Ghana [15, 16, 22–24].

## Results

### Model validation

Model validation results for the no-vaccination scenario across LMICs in 2016 among children under 5 years of age are presented in S6 Fig. There was strong agreement between model-predicted and proxy-estimated RVGE cases, with a Spearman correlation coefficient of 0.94. However, the model slightly underestimated the number of cases relative to the proxy data.

### Historical impact of rotavirus vaccination

Under the historical rotavirus vaccination program, the model estimated a median of 2.1 billion RVGE cases (95% uncertainty interval [UI]: 1.7 to 2.6 billion) across the 112 LMICs between 2006 (the year of first vaccine introduction) and the end of 2024 (Table 1). The estimated median annual RVGE cases declined substantially over time, from 120 million cases (interquartile range [IQR]: 112 to 130 million) in 2006, when only three countries had introduced the vaccine, to 85 million cases (IQR: 80 to 91 million) in 2024, by which time 81 countries had implemented vaccination programs (Fig 2A). This decline corresponds with the increasing number of countries adopting the vaccine, particularly from 2016 onward, when more than half of the 112 LMICs had introduced it (Fig 2A). In a counterfactual scenario with no vaccine introduction in any country, the model projected a median of 2.4 billion RVGE cases (95% UI: 1.9 to 2.9 billion) across the same 19-year period. In this no-vaccine scenario, annual RVGE cases were estimated at 120 million (IQR: 112 to 130 million) in 2006 and 122 million (IQR: 115 to 131 million) in 2024. Overall, the model estimated a median of 268 million averted RVGE cases (95% UI: 228 to 306 million) between 2006 and 2024. The annual number of averted cases increased substantially over time, from 37,000 (95% UI: 20,000 to 61,000) in 2006 to 37 million (95% UI: 31 to 44 million) in 2024.

**Fig 2.**
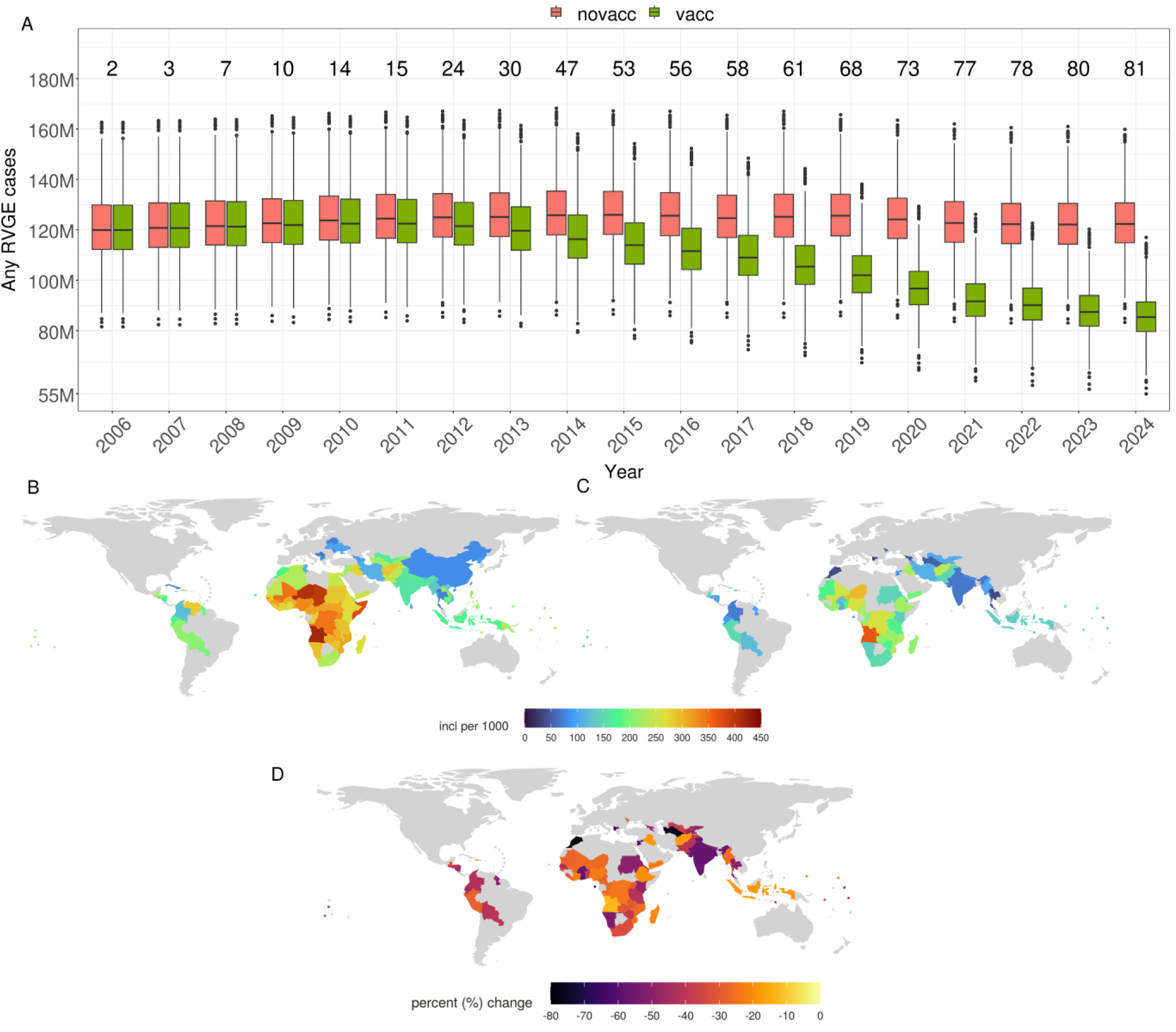
Impact of the historical rotavirus vaccination program across 112 LMICs from 2006 to 2024. (A) Box plots of annual model-simulated RVGE cases with (green) and without (red) vaccination across all LMICs. Values above the plots indicate the cumulative number of countries that introduced the vaccine by each year. Spatial distribution of model-predicted RVGE incidence (per 1,000 person-years) in (B) the absence of vaccination and (C) with vaccination for countries that have introduced the vaccine. (D) Percent reduction in RVGE incidence due to vaccination among countries that implemented the vaccine. For each country, the percent change is calculated from the year of vaccine introduction through the end of 2024.

**Table 1.**
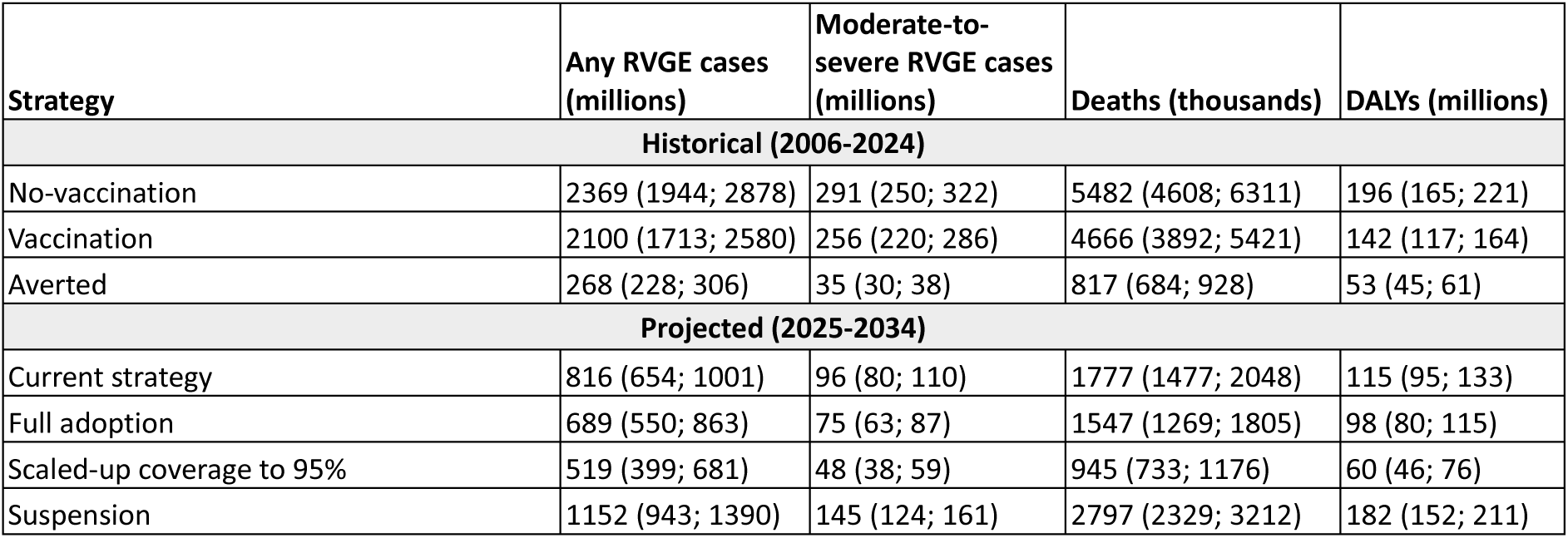
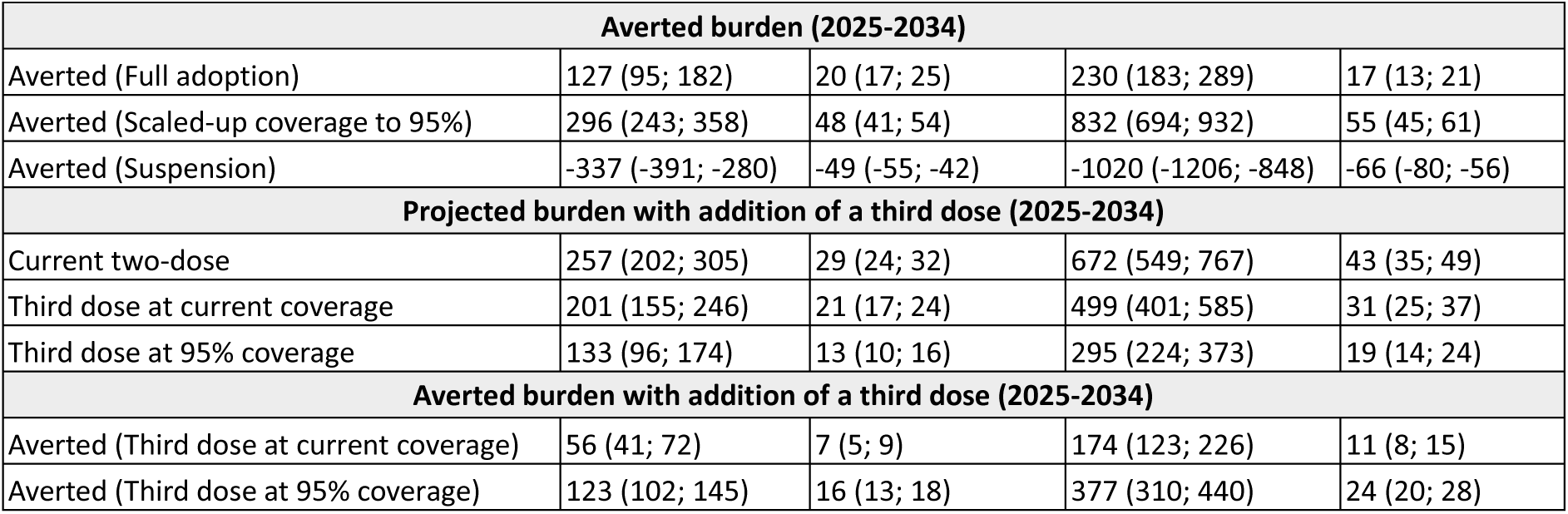
Summary of model-estimated historical and projected health outcomes: cases, deaths, and disability-adjusted life years (DALYs) for various vaccination strategies. Averted health outcomes are estimated using the no-vaccination scenario as the baseline for historical outcomes, the current vaccination program as the baseline for projected outcomes, and the current two-dose schedule as the baseline for the addition of a third-dose scenario. Values in parentheses represent interquartile ranges.

| Strategy | Any RVGE cases (millions) | Moderate-to-severe RVGE cases (millions) | Deaths (thousands) | DALYs (millions) |
| --- | --- | --- | --- | --- |
| **Historical (2006-2024)** |  |  |  |  |
| No-vaccination | 2369 (1944; 2878) | 291 (250; 322) | 5482 (4608; 6311) | 196 (165; 221) |
| Vaccination | 2100 (1713; 2580) | 256 (220; 286) | 4666 (3892; 5421) | 142 (117; 164) |
| Averted | 268 (228; 306) | 35 (30; 38) | 817 (684; 928) | 53 (45; 61) |
| **Projected (2025-2034)** |  |  |  |  |
| Current strategy | 816 (654; 1001) | 96 (80; 110) | 1777 (1477; 2048) | 115 (95; 133) |
| Full adoption | 689 (550; 863) | 75 (63; 87) | 1547 (1269; 1805) | 98 (80; 115) |
| Scaled-up coverage to 95% | 519 (399; 681) | 48 (38; 59) | 945 (733; 1176) | 60 (46; 76) |
| Suspension | 1152 (943; 1390) | 145 (124; 161) | 2797 (2329; 3212) | 182 (152; 211) |

| Averted burden (2025-2034) |  |  |  |  |
| --- | --- | --- | --- | --- |
| Averted (Full adoption) | 127 (95; 182) | 20 (17; 25) | 230 (183; 289) | 17 (13; 21) |
| Averted (Scaled-up coverage to 95%) | 296 (243; 358) | 48 (41; 54) | 832 (694; 932) | 55 (45; 61) |
| Averted (Suspension) | -337 (-391; -280) | -49 (-55; -42) | -1020 (-1206; -848) | -66 (-80; -56) |
| Projected burden with addition of a third dose (2025-2034) |  |  |  |  |
| Current two-dose | 257 (202; 305) | 29 (24; 32) | 672 (549; 767) | 43 (35; 49) |
| Third dose at current coverage | 201 (155; 246) | 21 (17; 24) | 499 (401; 585) | 31 (25; 37) |
| Third dose at 95% coverage | 133 (96; 174) | 13 (10; 16) | 295 (224; 373) | 19 (14; 24) |
| Averted burden with addition of a third dose (2025-2034) |  |  |  |  |
| Averted (Third dose at current coverage) | 56 (41; 72) | 7 (5; 9) | 174 (123; 226) | 11 (8; 15) |
| Averted (Third dose at 95% coverage) | 123 (102; 145) | 16 (13; 18) | 377 (310; 440) | 24 (20; 28) |

In the no-vaccination scenario, the model estimated a median RVGE incidence of 237 cases per 1,000 person-years (IQR: 229 to 245) across the 112 LMICs between 2006 and 2024 (Fig 2B). The highest country-specific incidence was predicted in Angola, with a median of 418 cases per 1,000 person-years (IQR: 337 to 454), while the lowest was in Serbia, with 68 cases per 1,000 person-years (IQR: 65 to 100). There was substantial heterogeneity in incidence across countries (S2 Appendix), with the highest burden estimated in those within the WHO African Region (Fig 2B). The spatial patterns closely follow the distribution of country-specific transmission rates (*R_0_*) and crude birth rates (S2 and S4 Fig).

Among the 81 countries that had implemented rotavirus vaccination by the end of 2024, the model estimated a median overall RVGE incidence of 205 cases per 1,000 person-years (IQR: 198 to 213). Country-specific incidence varied widely, ranging from 21 cases per 1,000 person-years (IQR: 19 to 24) in Albania to 364 cases per 1,000 person-years (IQR: 294 to 399) in Angola (Fig 2C). There was considerable heterogeneity in RVGE incidence across vaccinated countries (S2 Appendix), reflecting differences in time since vaccine introduction, coverage levels, and underlying demographic and transmission dynamics (S1 and S7 Fig).

The median overall impact of rotavirus vaccination in countries that introduced the vaccine, quantified as the percent reduction in RVGE cases compared to a no-vaccination baseline, was 34% (IQR: 31% to 38%). The highest country-specific median percent reduction was observed in Morocco at 79% (IQR: 70% to 85%), while the lowest was in Angola at 13% (IQR: 12% to 14%) (Fig 2D). Fifteen countries achieved a reduction of more than 50% in RVGE cases following vaccine introduction (S2 Appendix). Countries with the lowest estimated reductions were primarily those that introduced the vaccine more recently and generally had lower vaccination coverage (see S7 Fig).

The trends in moderate-to-severe RVGE cases, deaths, and DALYs followed a pattern similar to that of RVGE cases of any severity (S8 Fig). The historical vaccination program is estimated to have averted a median of 35 million moderate-to-severe RVGE cases (95% UI: 30 to 38 million), 817 thousand deaths (95% UI: 684 to 928 thousand), and 53 million DALYs (95% UI: 45 to 61 million) among children <5 years of age (Table 1). The averted DALYs were substantially higher in children <1 year, with 50 million DALYs (95% UI: 42 to 60 million), compared to 3 million DALYs (95% UI: 0 to 6 million) in those aged 1 to 4 years (S3 Table). The patterns of averted DALYs are consistent across WHO regions, with the highest and lowest percent of averted DALYs estimated in the South-East Asia Region and the African Region, respectively (S9 Fig). There is also considerable variation in the averted DALYs across countries with the vaccine (S2 Appendix).

### Projected vaccination scenarios between 2025 and 2034

The model projects substantial variation in any RVGE cases across 112 LMICs under different vaccination strategies over a 10-year period (Fig 3A). The impact of these strategies varies across the countries (S10 Fig and S3 Appendix). Under the status quo (i.e., countries maintaining current programs), the median estimated number of RVGE cases is 816 million (95% UI: 654 to 1001 million) (Table 1). In the full adoption scenario, where 29 non-vaccinating countries introduce the vaccine using a 6/10/14-week schedule at existing DTP3 coverage, the median decreases to 689 million (95% UI: 550 to 863 million). With coverage scaled up to 95% in countries currently below that threshold, the median estimated RVGE cases further decline to 519 million (95% UI: 399 to 681 million). Compared to the current strategy scenario, the full adoption scenario is projected to avert a median of 127 million RVGE cases (95% UI: 95 to 182 million), while the scaled-up coverage to 95% scenario would avert an estimated 296 million cases (95% UI: 243 to 358 million) over the 10-year period (Table 1). If vaccination programs were to be suspended across all 112 LMICs, the median RVGE cases are predicted to increase substantially to 337 million (95% UI: 280 to 391 million).

**Fig 3.**
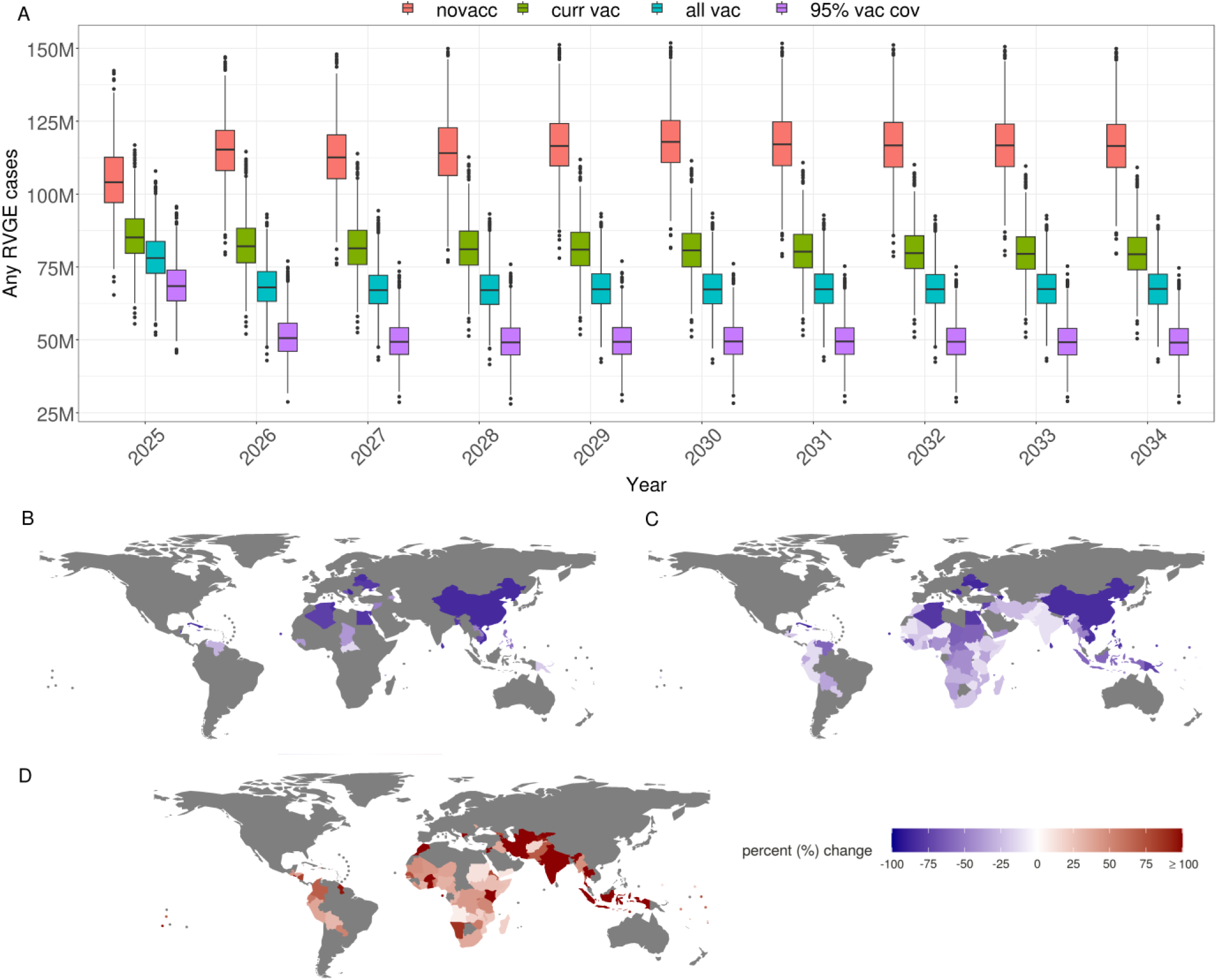
Projected impact of rotavirus vaccination on RVGE cases among children <5 years of age across 112 LMICs over a 10-year period (2025–2034). (A) Box plots of annual model-predicted RVGE cases under four vaccination scenarios: suspension or no introduction of vaccination in non-vaccine countries (novacc; red), continuation of current vaccination programs (curr vac; green), introduction of a 3-dose (6/10/14-week schedule) vaccine in currently unvaccinated countries at their existing DTP3 coverage levels (all vac; blue), and scaling up coverage to 95% (95% vac cov; purple) in countries with rotavirus or DTP3 coverage below 95% (countries with coverage ≥95% maintain their current coverage). (B) Percent change in RVGE cases relative to the current vaccination scenario following vaccine introduction in 29 countries without it. (C) Percent change in RVGE cases resulting from 95% scaled-up coverage. (D) Percent change in RVGE cases due to suspension or no vaccine introduction in countries without vaccination.

The model projects a wide range of percentage changes in RVGE cases across the vaccination scenarios, using current country-specific vaccination programs as the baseline (Fig 3B-D). Under the full adoption scenario, in which the 29 non-vaccinating countries introduce vaccination with a 6/10/14-week schedule at existing DTP3 coverage, overall RVGE cases are projected to decline by a median of 15% (IQR: 13%–17%), ranging from 14% (IQR: 13%–16%) in the Central African Republic to 87% (IQR: 86%–88%) in Cuba (Fig 3B). The model projected an overall median percent change under the scaled-up coverage to 95% scenario to be 36% (IQR: 34% to 38%), with a greater reduction estimated in countries with low current rotavirus or DTP3 coverage (Fig 3C; see S1 Fig). In contrast, suspending vaccination programs in the 83 currently vaccinating countries is projected to result in a median 41% (IQR: 39% to 44%) overall increase in RVGE cases compared to current vaccination (Fig 3D). The model projected a substantial increase in RVGE cases of more than 100% for about 20 countries (S3 Appendix).

There are substantial differences in the projected impact of these strategies on moderate-to-severe RVGE cases, deaths, and DALYs (S11 Fig). The trends in the differential percent of DALYs averted for these strategies are consistent, although the absolute impact varies across WHO regions (S12 Fig). Detailed country-specific estimates of cases, deaths, and DALYs for each scenario are presented in S3 Appendix.

In the 29 countries without rotavirus vaccination programs, similar trends in projected RVGE cases were observed for the 6/10- and 10/14-week schedules (S13 Fig), with the 6/10-week schedule performing slightly better. Across WHO regions, using no vaccination as the baseline, the 6/10-week schedule consistently predicted a higher percent of DALYs averted (S14 Fig). However, the impact of the two-dose schedules was still lower compared to the three-dose 6/10/14-week schedule.

### Adding an additional dose to the two-dose vaccination schedule

For the 51 countries currently using the standard two-dose rotavirus vaccination schedule, the impact of adding a third dose varied across countries (Fig 4). Compared to the two-dose schedule, the overall median number of additional averted RVGE cases was 56 million (95% UI: 41 to 72 million), corresponding to a 22% (IQR: 20% to 24%) reduction at current coverage levels over the 10 years. At 95% coverage, averted cases increased to 123 million (95% UI: 102 to 145 million), with a median reduction of 48% (IQR: 46% to 51%). Despite this, the country-specific impact of an additional dose was moderate at the current coverage but substantial at the 95% scaled-up coverage (Fig 4B and C). The country-specific incidence is provided in S15 Fig and S4 Appendix.

**Fig 4.**
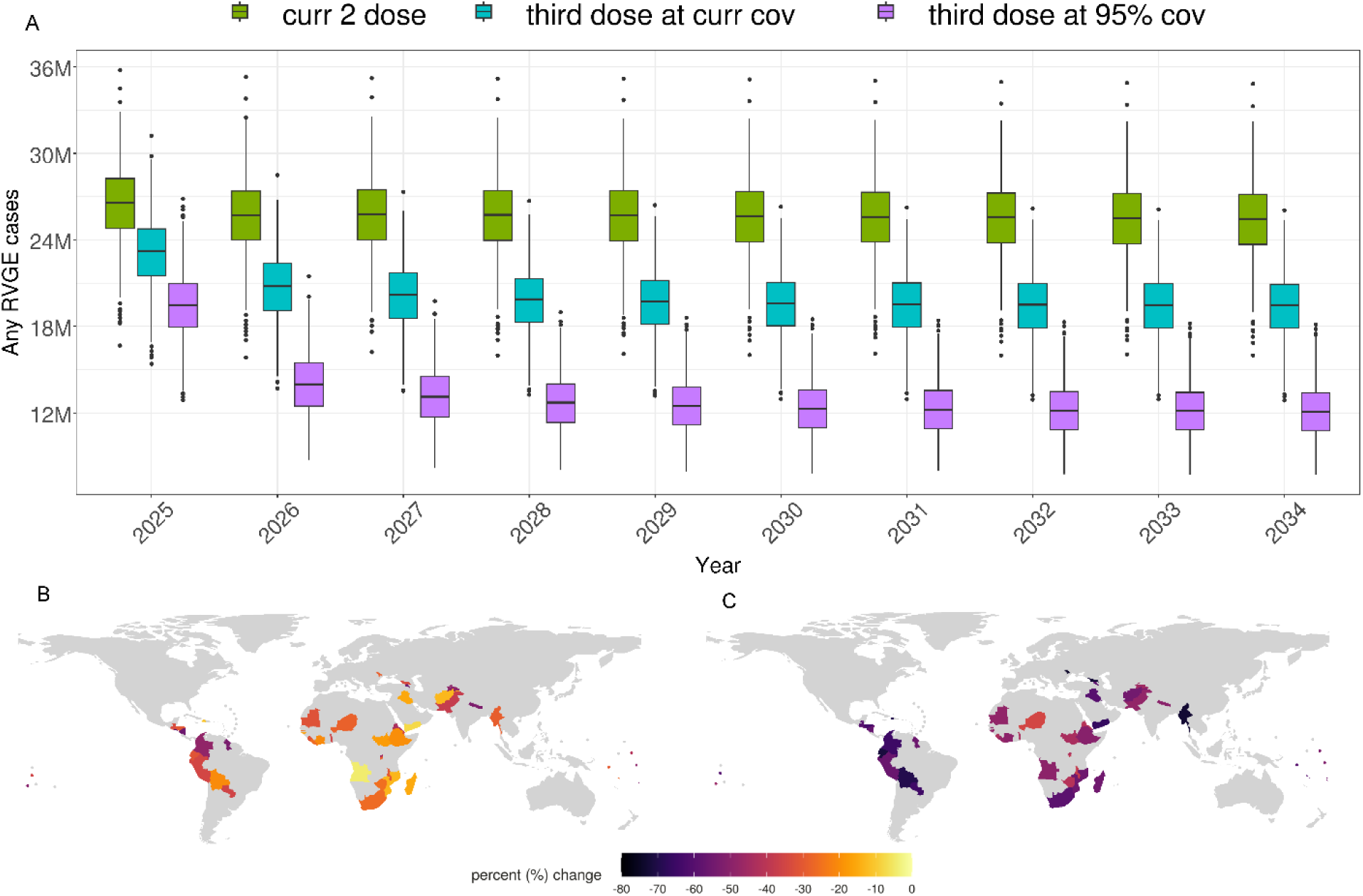
Model-projected impact of adding a third rotavirus vaccine dose in the 51 countries currently using a two-dose schedule (2025–2034). (A) Box plots of annual model-simulated RVGE cases under three scenarios: continuation of the current two-dose schedule (green), addition of a third dose at current vaccination coverage levels (blue), and addition of a third dose with coverage scaled up to 95% (countries already above 95% maintain their current coverage; pink). (B) Percent change in RVGE cases relative to the current two-dose schedule and the scenario with an additional dose at current coverage. (C) Percent change in RVGE cases relative to the current two-dose schedule and the scenario with an additional dose at 95% scaled-up coverage.

Adding a third vaccine dose in countries currently implementing a two-dose schedule increased the estimated health impact, although the most substantial benefits were observed under the 95% scaled-up coverage scenario (S16 Fig, S3 Table). Compared with the two-dose schedule, the projected median averted moderate-to-severe RVGE cases was 7 million (95% UI: 5 to 9 million), 174 thousand (95% UI: 123 to 226 thousand) deaths, and 11 million (95% UI: 8 to 15 million) DALYs (Table 1). Under the 95% scaled-up coverage scenario, the projected health impact was markedly greater, with an estimated 16 million (95% UI: 13 to 18 million) cases, 377 thousand (95% UI: 310 to 440 thousand) deaths, and 24 million (95% UI: 20 to 28 million) DALYs averted. Across the WHO regions, with the exception of South-East Asia Region, adding a third dose at current coverage resulted in a higher median percentage of DALYs averted among children aged 1–4 years compared with those <1 year (S17 Fig). However, when the third dose was added at 95% scaled-up coverage, the median percentage of DALYs averted was higher in the <1 year age group in all regions except European Region. Despite this, there are substantial variations in the health impact of the additional dose at the country level (S4 Appendix).

## Discussion

We provide robust estimates of the health impact of historical rotavirus vaccination programs and quantify the potential benefits of alternative strategies to improve their effectiveness across 112 LMICs. Strategies such as scaling up coverage in countries with existing vaccination programs, implementing high-coverage vaccination programs in countries without existing vaccination, and adding a third dose to the standard two-dose schedule could significantly enhance rotavirus vaccine impact and lead to substantial reductions in cases, deaths, and DALYs compared to the current program. Our findings highlight the significant public health benefits of rotavirus vaccination, supporting its continued sustainability and expansion. This modeling framework provides a valuable tool for country-specific evaluations of rotavirus immunization programs and can inform policy decisions on the introduction, evaluation, and optimization of vaccination strategies across LMICs.

A key strength of this modeling study lies in its robust estimation of the historical impact of rotavirus vaccination and its evaluation of the relative effectiveness of alternative immunization strategies compared to the current vaccination program. By assessing commonly implemented infant rotavirus vaccination schedules, this analysis provides valuable country-specific insights for nations considering the introduction or optimization of vaccination programs. This comprehensive modeling framework distinguishes our approach from previous studies that primarily evaluate existing programs or single strategies against a no-vaccination baseline [18, 20, 25]. For resource-limited countries that have not yet introduced rotavirus vaccines and often lack surveillance data, our model provides critical evidence on both the existing disease burden and the potential health benefits of vaccination. Furthermore, the use of a model that has demonstrated strong performance during extensive validation adds credibility to our results. For instance, the model successfully reproduced observed pre-vaccination rotavirus trends in Bangladesh and cases before and after vaccine introduction in Ghana and Malawi [15, 16, 21–24]. It also realistically simulated vaccine impacts under conditions such as COVID-19–related disruptions and vaccine product transitions [23, 24], demonstrating its robustness. A strong correlation between our model projections and regression-based estimates of rotavirus burden across LMICs from the Global Burden of Disease study (S6 Fig) further supports the predictive validity of our approach [32]. Additionally, the no-vaccination estimates align well with pre-introduction burden estimates, reinforcing the reliability of our findings [33].

Our results indicate a substantial overall impact of rotavirus vaccination across the 81 countries that had introduced the vaccine by the end of 2024. However, the estimated vaccine impact varied considerably between countries. These differences can be attributed to several factors, including background RVGE transmission dynamics, birth rate, country-specific variation in vaccine response, coverage levels, and duration since vaccine introduction. For example, Angola showed a lower estimated impact due to its high birth rate, higher transmission rate, reduced vaccine response, and lower vaccine coverage. Among countries that introduced the vaccine between 2020 and 2024, the overall reduction in RVGE was moderate, ranging from 9% to 43% (S2 Appendix). In contrast, countries with rotavirus vaccination programs for more than a decade and higher vaccination coverage have experienced reductions of 37% to 79% (S2 Appendix). These findings are consistent with observational studies [7, 34, 35], which demonstrate that vaccine impact increases over time after introduction. This time-dependent improvement in population-level protection is closely linked to increasing vaccination coverage and may partly reflect the accumulation of indirect (herd) effects as sustained vaccination reduces community transmission. Although rotavirus vaccines provide some herd immunity in LMICs, this effect remains heterogeneous across settings [19,36–38]. Expanding vaccine coverage could improve indirect protection and further reduce the burden of RVGE. This is particularly relevant in LMICs, where high transmission intensity and birth rates can limit the effectiveness of low-coverage vaccination programs. Scaling up vaccination coverage across LMICs, as illustrated in our 95% scaled-up coverage scenario projections, is therefore essential to maximize both the direct and indirect benefits of rotavirus vaccination.

An additional dose to the standard two-dose Rotarix schedule, as well as altering dosing schedules, has been suggested as a potential strategy to improve vaccine impact in LMICs [14, 16, 39–41]. However, immunogenicity and efficacy estimates from clinical trials evaluating an additional dose of Rotarix have generally been comparable to, or only moderately higher than, the standard schedule across LMICs [12, 13, 41–43]. Nevertheless, adding an extra dose to the standard two-dose Rotarix schedule could enhance protection by providing earlier protection when administered at 6 weeks before the 10/14-week schedule begins or by extending protection when an additional dose is given at 14 weeks in the 6/10-week schedule. Overall, a third dose offers limited additional benefits unless vaccination coverage improves. At 95% coverage, the additional dose yields a substantial impact, although the magnitude of this benefit varies across countries. Thus, countries considering the addition of an extra dose should conduct country-specific impact analyses that account for vaccine coverage, rotavirus epidemiology, and the current impact of the two-dose schedule.

Across the 29 countries without rotavirus vaccination, we estimated similar but modestly higher DALYs averted under the 6/10-week schedule compared with the 10/14-week schedule. This difference in vaccine impact is likely due to the earlier first-dose administration in the 6/10-week schedule, which provides protection in LMICs where severe RVGE typically occurs early in life [44]. Additionally, infants exposed to RVGE before receiving their first vaccine dose have a lower probability of mounting an immune response [45, 46]; thus, earlier vaccination increases the likelihood of achieving an improved vaccine response. However, some studies suggest that higher maternal antibody levels at the time of vaccination can reduce vaccine immunogenicity [47, 48]. Together, these findings highlight that tailoring two-dose schedules to local epidemiological patterns could improve the health impact of rotavirus vaccination in LMICs.

Currently, limited evidence exists on the health impacts of suspending vaccination or the introduction of vaccines in countries without existing programs. This information is of critical public health importance, as rotavirus vaccination has already been suspended in the Philippines and Venezuela, and 29 countries out of the 112 LMICs have yet to introduce the vaccine. Funding for rotavirus vaccine programs is likely to be further constrained under the current global health climate. Using country-specific vaccination strategies as the baseline, our model projections indicate that discontinuation could lead to a substantial resurgence of RVGE burden, with cases increasing by more than 400% in some countries over a 10-year period. Conversely, introducing rotavirus vaccination in the 29 currently unvaccinated countries is projected to markedly reduce cases and deaths and improve overall population health, as demonstrated by substantial averted DALYs. These findings demonstrate rotavirus vaccination as an effective public health intervention and support the continuation and expansion of immunization programs across the 112 LMICs.

Several limitations of our study warrant consideration in the interpretation of our findings. First, we assumed equivalent vaccine efficacy across different rotavirus vaccines when estimating the vaccine response rates used in our model. While efficacy estimates from clinical trials across LMICs have shown generally comparable results, even within the same geographic settings, such as trials conducted in Ghana [49–51], real-world effectiveness may vary, potentially introducing bias into our estimates [24]. Second, our estimates of vaccine response rates are derived from a previously published regression-based model of rotavirus vaccine efficacy [11], which may not fully capture country-specific variations in immune response. Third, we applied the same duration of vaccine-induced protection across all countries. While we sampled from the range of protection durations estimated when our model was fitted separately to surveillance data from Ghana and Malawi for both Rotarix and Rotavac vaccines [15, 16, 22, 24], this approach may not fully capture the potential heterogeneity in waning vaccine-induced immunity across different settings. We estimated substantial variation within and across countries in the duration of vaccine-derived immunity for the Rotarix vaccine [15, 22]. Fourth, in projecting vaccine impact over a 10-year period, we assumed fixed vaccination coverage levels based either on current rotavirus or DTP3 coverage. While this assumption may yield conservative estimates for recently introduced countries (where coverage is likely to increase over time), it may overestimate the impact for countries where coverage remains persistently low. Although we partially addressed the coverage issue by running a scenario with fixed 95% coverage, we are likely overestimating the impacts, as coverage remains below 70% in some countries even after more than a decade of implementation [52]. Future work incorporating country-specific calibration of model parameters will be essential to improve the robustness of projected vaccine impact.

Our modeling approach can generate evidence essential for guiding immunization policy. Importantly, our results support WHO recommendations for the universal introduction of rotavirus vaccines, particularly in LMICs where the burden remains high. By quantifying the impact of current programs and evaluating alternative implementation strategies, our framework shows how RVGE burden could be further reduced through optimized vaccination strategies. Although this analysis focuses on health outcomes, the country-specific burden estimates generated by the model can readily support comparative economic evaluations needed to identify the most cost-effective strategies for each country. Overall, this modeling framework provides a practical decision-support tool for addressing key policy questions, including the current impact of vaccination and the potential benefits of alternative strategies. It can support national decision-makers in identifying optimal, evidence-based approaches that maximize the public health impact of rotavirus vaccination across LMICs.

## Data Availability

All data produced in the present work are contained in the manuscript

## Acknowledgements

Nigel Cunliffe is a National Institute for Health and Care Research (NIHR) Senior Investigator (NIHR203756). The views expressed in this article are those of the author(s) and not necessarily those of the NIHR, or the Department of Health and Social Care.

## Competing interests

VEP has received funding from Merck for an investigator-initiated grant unrelated to the topic of this manuscript. All other authors declare no competing interests.

## Notes

### Funding Statement

National Institutes of Health grant R01AI112970 (VEP)

