## Supplementary material for "Quantifying and identifying strategies to improve rotavirus vaccine impact in low- and middle-income countries": S1 Text.pdf

**Short title: Optimizing rotavirus vaccines in LMICs**

Ernest O. Asare[1,2*], Jiye Kwon[1,2], Xiao Li[1,3], Mohammad A. Al-Mamun[4], Belinda L. Lartey[5], Khuzwayo C. Jere[6,7,8], Nigel A. Cunliffe[6], George E. Armah[5], Benjamin A. Lopman[7], Virginia E. Pitzer[1,2]

**The PDF file includes:**

Materials and Methods
S1 Fig to S17 Fig
S1 Table to S3 Table
References

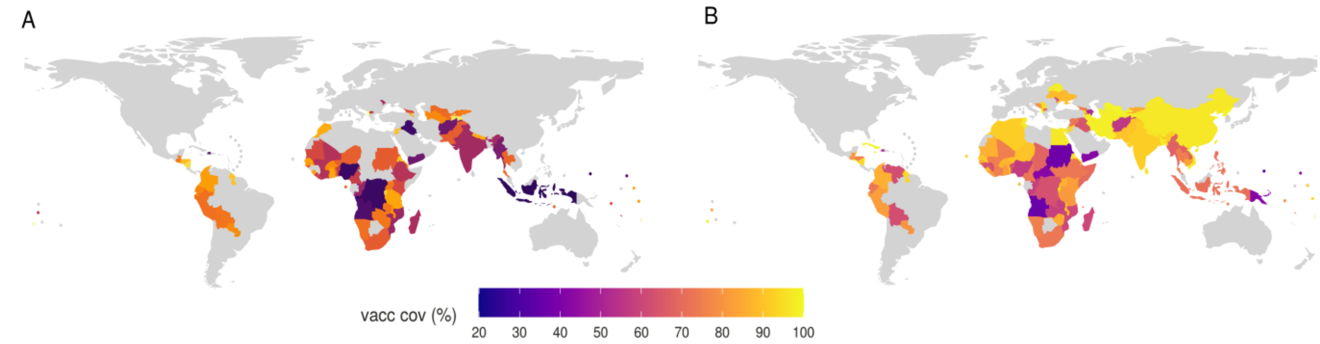


**S1 Fig. Country-specific vaccination coverage.** (A) Average rotavirus vaccination coverage for each country from the year of vaccine introduction through 2024, used in retrospective simulations. (B) Rotavirus vaccination coverage in 2024 for countries that have introduced the vaccine, and DTP3 coverage in 2024 for countries that have not. Data sourced from the WHO/UNICEF Estimates of National Immunization Coverage (WUENIC) [1].

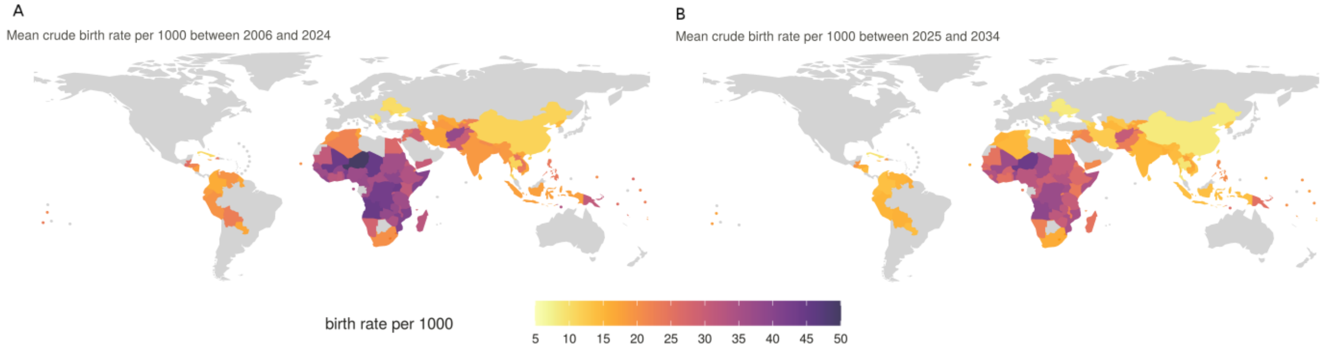


**S2 Fig. Average country-specific crude birth rates per 1000 for 112 LMICs.** (A) Average rates from 2006 to 2024, used in retrospective simulations. (B) Average rates from 2025 to 2034, used in projection simulations. Data sourced from the U.S. Census Bureau [2].

***Model structure description***

We applied a deterministic, age-stratified compartmental model of rotavirus transmission (S3 Fig) to estimate the historical and projected impact of the rotavirus vaccination program across 112 low- and middle-income countries (LMICs). This model has been previously developed, calibrated, and validated against observed epidemiological data from three countries within the 112 LMICs [3–8]. The model was adapted for each country using country-specific model input data.

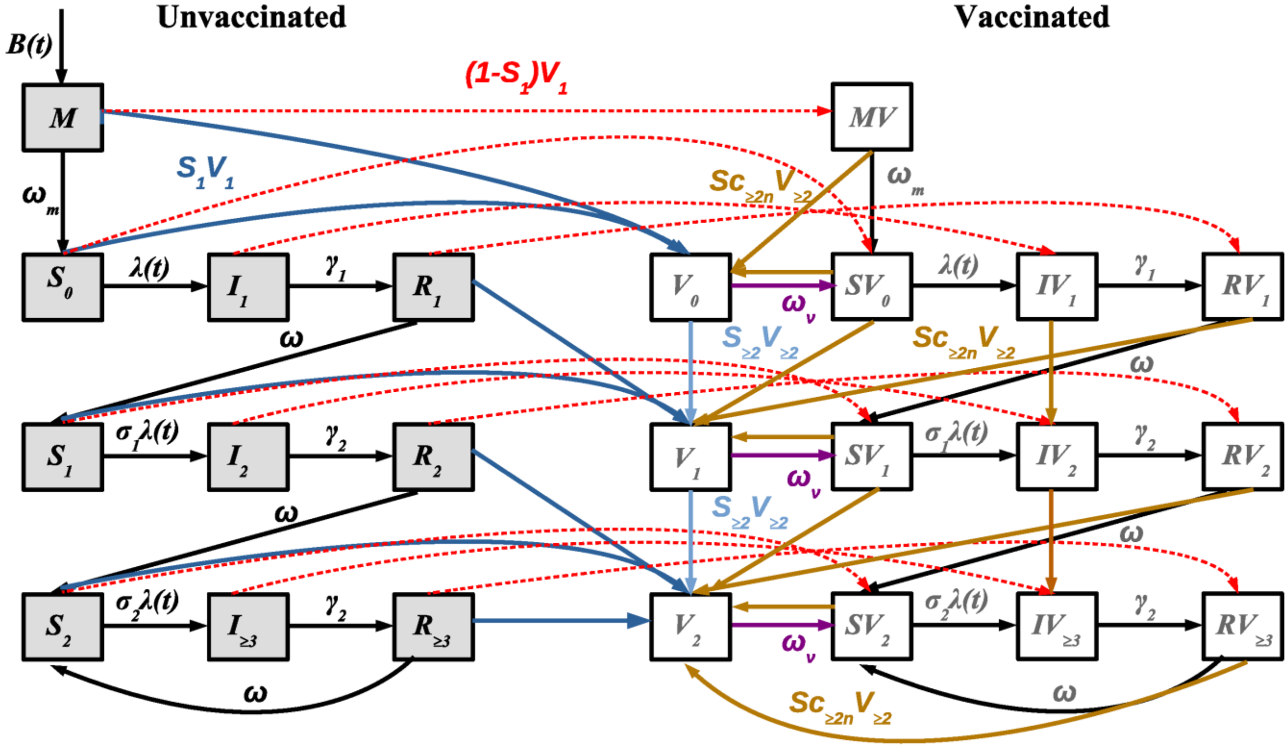


**S3 Fig. Schematic representation of the rotavirus transmission model**. Unvaccinated and vaccinated individuals are depicted by open and grey boxes, respectively. The compartments labeled *M*, *S*, *I*, and *R* correspond to maternal immunity, susceptible, infected, and recovered states. Arrows illustrate the directions in which individuals transition between compartments. Red lines denote the movement of individuals who did not respond to the first vaccine dose. Dark and light blue lines represent transitions of those who responded to the initial and subsequent doses, respectively. Gold lines indicate transitions based on the likelihood of responding to additional doses after an initial failure. Purple lines show transitions resulting from waning vaccine-induced immunity. The values of the fixed model parameters are defined in S1 Table.

**S1 Table. Fixed model parameters and their defined values**.

| Fixed parameters | Symbol | Value | Source |
|---|---|---|---|
| Relative risk of second infection | $\sigma_1$ | 0.62 | [9, 10] |
| Relative risk of third infection | $\sigma_2$ | 0.35 | [9, 10] |
| Relative infectiousness of secondary infection | $\rho_2$ | 0.5 | [11] |
| Relative infectiousness of mild/asymptomatic infections | $\rho_{\geq 2}$ | 0.1 | [11] |
| Duration of primary infection | $1/\gamma_1$ | 1 week | [12] |
| Duration of secondary infection | $1/\gamma_2$ | 0.5 week | [13, 14] |
| Duration of temporary immunity following infection | $1/\omega$ | 13 weeks | [15] |

Individuals enter the model at birth into a maternally protected compartment ($M$), at a rate corresponding to each country's birth rate ($B$). These infants are assumed to be temporarily protected from infection due to maternally acquired antibodies, which wane at a rate $\omega_m$. Once maternally derived immunity wanes, individuals transition to the fully susceptible compartment ($S_0$), where they are at risk of experiencing a primary rotavirus infection at a rate $\lambda$. Infected individuals enter the primary infection compartment ($I_1$), where a proportion ($d_1$) develop moderate-to-severe RVGE and remain infectious for an average duration of $1/\gamma_1$. Following recovery, individuals move to the first recovered compartment ($R_1$), where they are temporarily protected against reinfection for an average period of $1/\omega$. Once immunity from the primary infection wanes, individuals become susceptible to a second infection ($S_1$), but at a reduced transmission rate ($\sigma_1\lambda$). These secondary infections ($I_2$) are assumed to be less infectious (by a factor $\rho_2$), shorter in duration ($1/\gamma_2$), and less likely to result in moderate-to-severe RVGE ($d_2$) compared to primary infections. Recovered individuals from this stage enter the $R_2$ compartment, where they again gain temporary immunity that wanes at the same rate $\omega$. Subsequent waning of immunity leads individuals into a partially immune susceptible compartment ($S_2$), where subsequent infections can occur at a further reduced rate ($\sigma_2\lambda$). These subsequent infections ($I_{\geq 3}$) are typically mild or asymptomatic ($d_3$), and the infectiousness is further reduced (by a factor $\rho_{\geq 3}$). Recovery from these infections occurs at the same rate as for secondary infections ($1/\gamma_2$), after which individuals move into a temporary immune state ($R \geq 3$). When this immunity wanes ($1/\omega$), individuals return to the partially immune susceptible state ($S_2$), allowing for repeated cycles of mild or asymptomatic reinfection.

Vaccination was implemented by treating each dose as conferring immunity equivalent to one natural rotavirus infection among individuals who mount an immune response. Separate compartments were used to track vaccinated individuals, with the rate of transfer from the unvaccinated to the vaccinated compartment determined by country-specific first-dose vaccine coverage for each country. For individuals who do seroconvert, vaccination provides temporary protection against rotavirus infection, denoted by the $V_i$ compartment, which waned over time at a rate denoted by $\omega_v$ (same for all doses). Once vaccine-induced immunity waned, individuals became susceptible to subsequent infections: primary infection after one dose, secondary after two doses, and tertiary after three doses. The model assumes heterogeneity in vaccine response: individuals who respond to the first dose have a higher probability of responding to subsequent doses compared to those who fail to respond to the first dose. Those who fail to respond to any dose remain in their current compartment, and their transitions between states follow those of the unvaccinated compartment. We used country-specific rotavirus vaccine schedules and year-specific coverage estimates from WHO/UNICEF for the current vaccine impact simulations. For projection simulations, we applied 2024 coverage levels based on either rotavirus vaccine or DTP3 coverage, depending on data availability. In countries without existing rotavirus vaccination programs, we assumed the use of standard two-dose and three-dose schedules (administered at 6/10, 10/14, or 6/10/14 weeks of age), consistent with schedules recommended for the four WHO-prequalified vaccines: Rotarix, RotaTeq, Rotavac, and Rotasiil. To align with routine immunization timelines, vaccination ages were approximated to the nearest month [16].

***Country-specific $R_0$ estimation***

For each country, we used country-specific demographic data (total population, crude birth rate, and crude death rate) to simulate a rotavirus transmission model across a wide range of transmission rate ($R_0$) values in the absence of vaccination. The simulations were run in finite steps across the $R_0$ range. We then compared the transmission model-estimated age of first severe rotavirus infection in 2015 with estimates obtained from a previously published regression-based approach [17]. For each country, we identified the $R_0$ values from our model that corresponded to an age of infection falling within the 95% confidence interval of the age estimated by the regression model. The mean estimate of $R_0$ for each country is shown in S4 Fig. To account for uncertainty in the estimated $R_0$ values, we assumed a normal distribution and performed 1,000 random samples between the minimum and maximum $R_0$ values obtained for each country. These sampled $R_0$ values were then used as input for the model simulations.

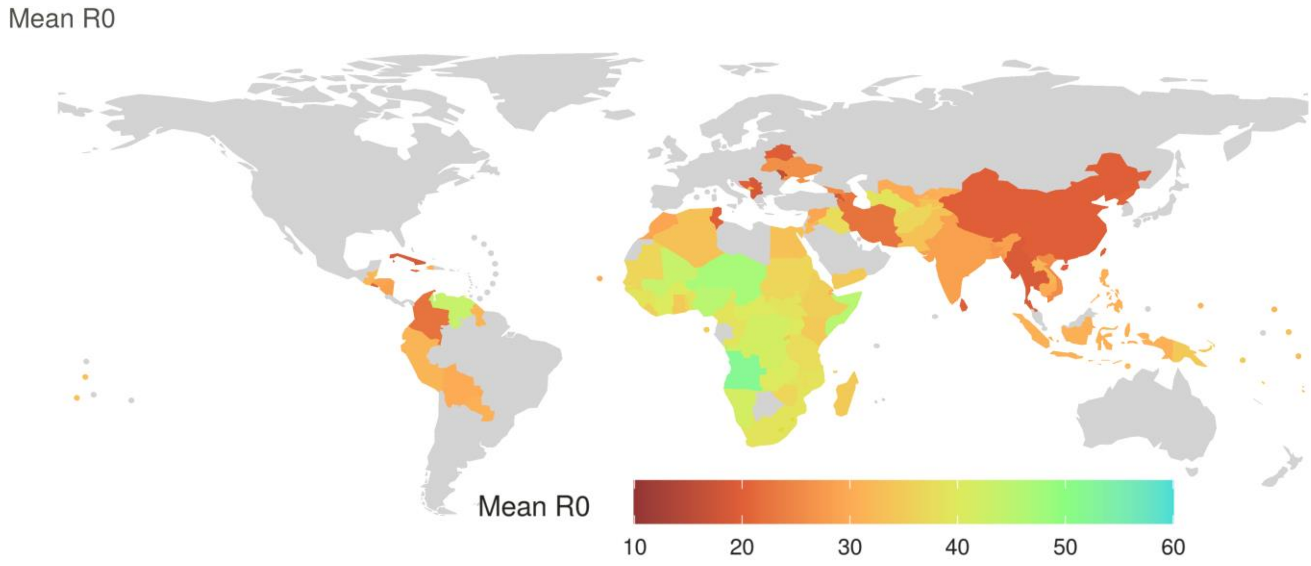


**S4 Fig. Estimated country-specific mean $R_0$ for 2015 across 112 LMICs**. The country-specific $R_0$ estimates and their 95% confidence intervals are provided in S1 Appendix.

***Seroconversion (vaccine response) estimate***

We estimated country-specific probabilities of vaccine response by calculating two key parameters, following the approach described by Pitzer et al. [3]: (1) the proportion of infants who are immunologically capable of responding to the vaccine (*m*, with $n = 1 - m$ representing non-responders), and (2) the probability of seroconversion among those capable of responding (*p*, with $q = 1 - p$ denoting non-response among responders).

The key assumption here is that there are infants that may not immune response regardless of the number of doses of the vaccine due to infant gut microbiome composition, malnutrition, and genetic factors [18–21]. Consequently, the population is divided into two groups:

Responders, with probability *m*

Non-responders, with probability $n = 1 - m$

Among responders, we define:

The probability of responding to a single vaccine dose as *p*

The probability of failing to respond as $q = 1 – p$

The probability of seroconversion after receiving two or three doses defined as follows:

After two doses:

Pr(Seroconversion|2 doses) = $p^2m$

After three doses:

Pr(Seroconversion|3 doses) = $(3p^2q + p^3)m$

From these expressions, we can calculate the probabilities of responding or not responding to each dose of the vaccine among responders (for full derivation of these equations, see [15]) as follows:

- The probability of responding to the first dose of the vaccine:

  $S_{C1} = pm$

- The probability of responding to the second dose, among those having responded to the first dose:

  $S_{C2} = p$

- The probability of responding to the third dose is equivalent to the probability of responding to the second dose:

  $S_{C3} = S_{C2}$

- The probability of responding to the second dose, given failure to respond to the first dose:

  $$S_{C2n} = \frac{qpm}{(1-pm)} = \frac{(1-S_{C2})S_{C1}}{(1-S_{C1})}$$

- For individuals who failed to respond to either of the first or second doses, the probability of seroconversion following a third dose is given by:

  $$S_{C3n} = \frac{(1-S_{C2})^2 S_{C1}}{1-m+m(1-S_{C2})^2}$$

***Efficacy and seroconversion relationship***

We utilized a simple linear regression model to estimate the relationship between vaccine response rate and vaccine efficacy. The model was fitted using published rotavirus clinical trials that reported both the response rate and efficacy of the four available rotavirus vaccines [22]. A separate regression model was fitted for two-dose schedules, three-dose schedules, and the combined data (i.e., both two- and three-dose schedules). Results of regression models are presented in S5 Fig.

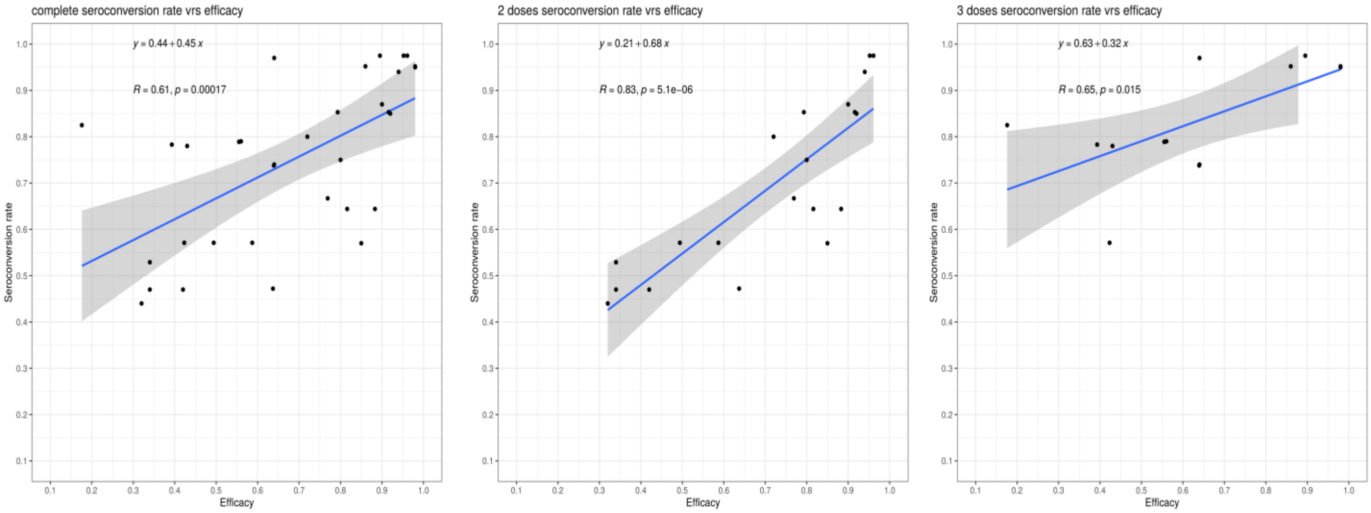


**S5 Fig. Results of the regression models assessing the association between rotavirus vaccine efficacy and response rate**. (A) Two-dose schedules, (A) three-dose schedules, and (C) combined two- and three-dose schedules. Data are derived from clinical trials of all four available rotavirus vaccines that reported both efficacy and immune response rates, as documented in reference [22].

### *Case fatality rate (CFR) and disability-adjusted life years (DALYs)*

Rotavirus-specific case fatality rates (CFR) were extracted from studies across various WHO regions from our previously published systematic review [23]. Due to the lack of sufficient rotavirus-specific CFR studies at the country level, we used WHO regions for the analysis, assuming that all countries within a given region would have the same estimated CFR. The pooled age-stratified CFR for each WHO region was estimated for two age groups: <1 year and 1-4 years, using a random-effects model, with 95% confidence intervals (CIs) calculated. To account for uncertainty in the CFR estimates, we generated 1,000 simulations by sampling uniformly within the 95% CIs for each region. The model-predicted incidence of moderate-to-severe RVGE for each age group was then multiplied by the corresponding CFR for that age group to estimate the mortality attributed to rotavirus for both the <1 year and 1-4 years age groups. The age-stratified CFR for each WHO region are summarized in S2 Table.

To approximate disability weights for moderate-to-severe rotavirus, we sampled from the lower bound of the 95% uncertainty interval for moderate diarrhea (0.125) and the upper bound of the 95% uncertainty interval for severe diarrhea (0.348), based on estimates from the Global Burden of Disease 2013 (24). We generated 1,000 samples from a beta distribution constrained to this range. The duration of illness was assumed to be 6 days (range: 5.9–6.1 days), based on published estimates [25, 26], with 1,000 samples generated from a normal distribution defined by this range.

**S2 Table. Estimated rotavirus-specific case fatality rates (CFRs, %) stratified by World Health Organization (WHO) region and age group**.

| **WHO Region** | **<1Y (95% CI)** | **1-4Y (95% CI)** |
|---|---|---|
| African Region (AFRO) | 1.535 (0.598-3.882) | 1.340 (0.444-3.974) |
| Region of the Americas (AMRO) | 0.765 (0.011-34.545) | 0.313 (0.020-4.665) |
| South-East Asia Region (SEARO) | 0.205 (0.008-4.806) | 0.211 (0.051-0.858) |
| European Region (EURO) | 0.093 (0.002-3.695) | 0.090 (0.034-0.239) |
| Eastern Mediterranean Region (EMRO) | 0.073 (0.000-17.260) | 0.104 (0.003-3.234) |
| Western Pacific Region (WPRO) | 0.036 (0.001-1.526) | 0.205 (0.058-0.713) |

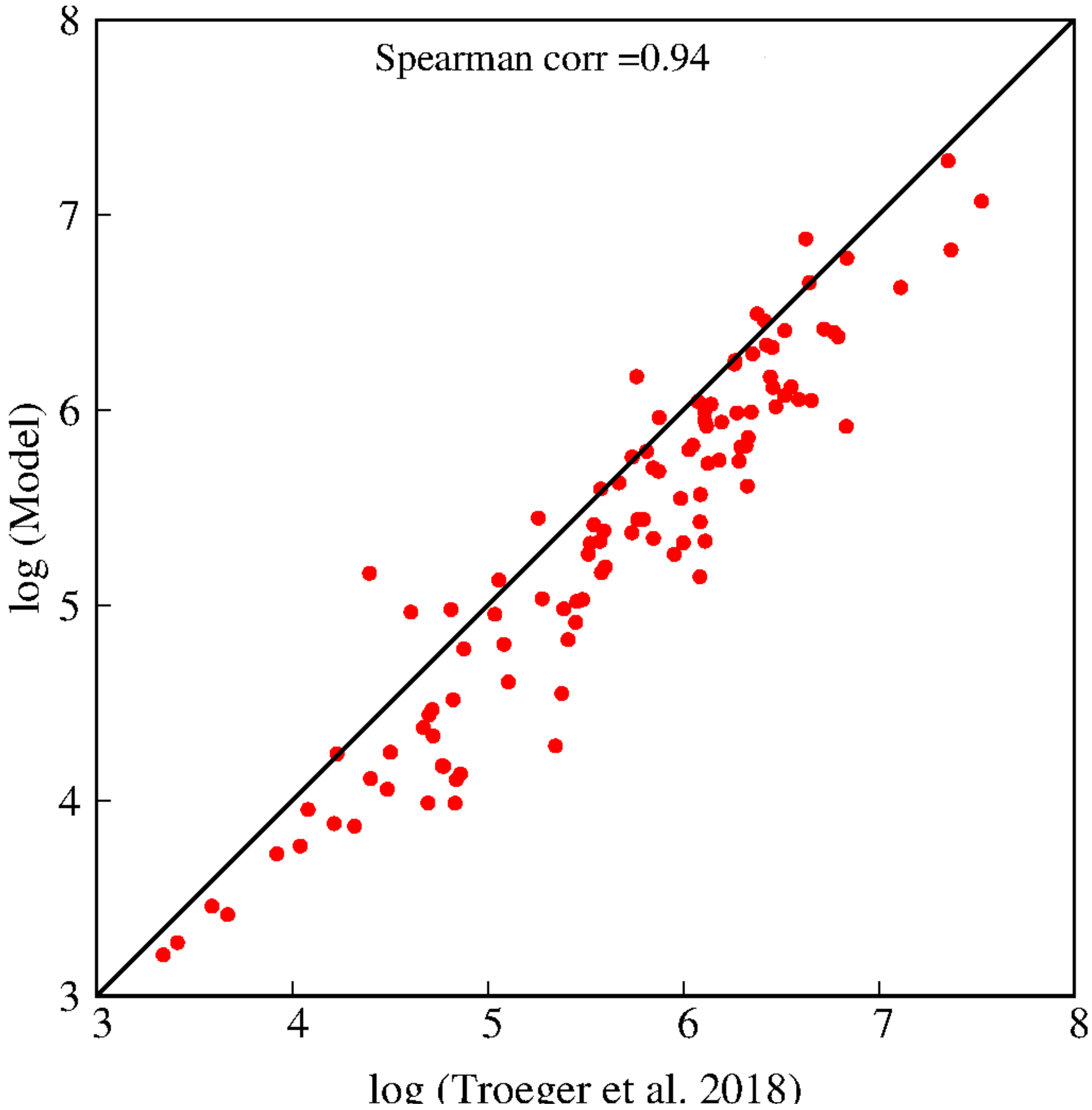


**S6 Fig. Scatter plot comparing model-estimated and proxy RVGE cases in children under 5 years of age in 2016 across LMICs, in the absence of vaccination**. Proxy estimates are taken from Troeger et al. 2018 [27]. Both model and proxy data have been log-transformed.

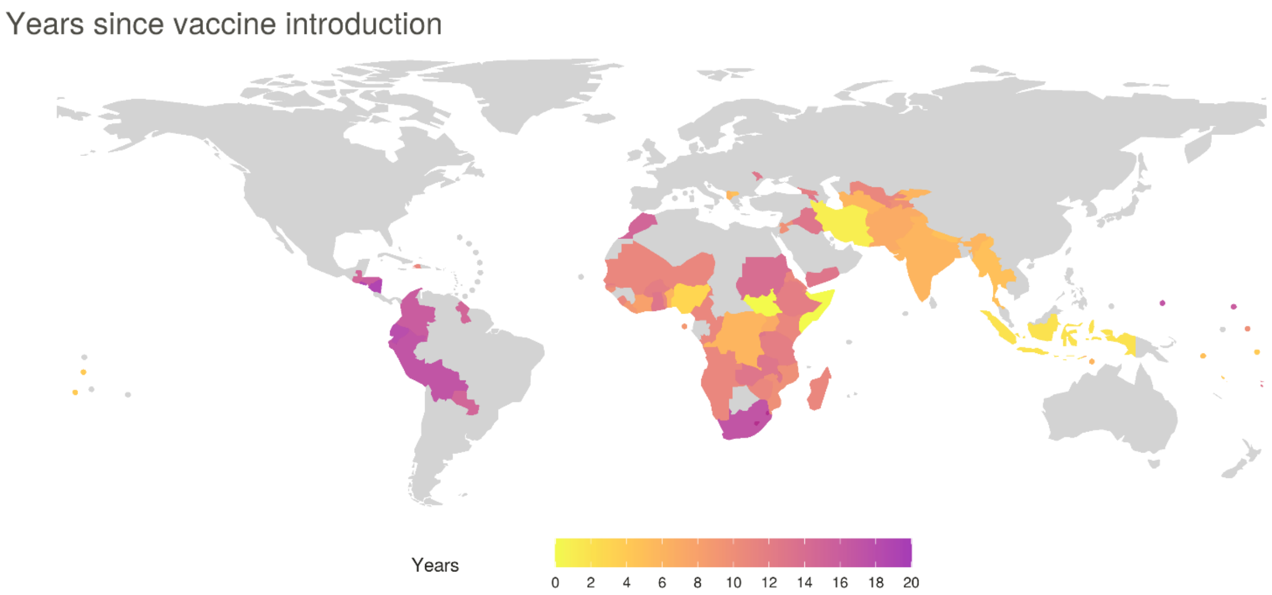


**S7 Fig. Spatial distribution of years since vaccine introduction across the 81 LMICs that had implemented vaccination by 2024**. For each country, years are calculated from the year of vaccine introduction through 2024.

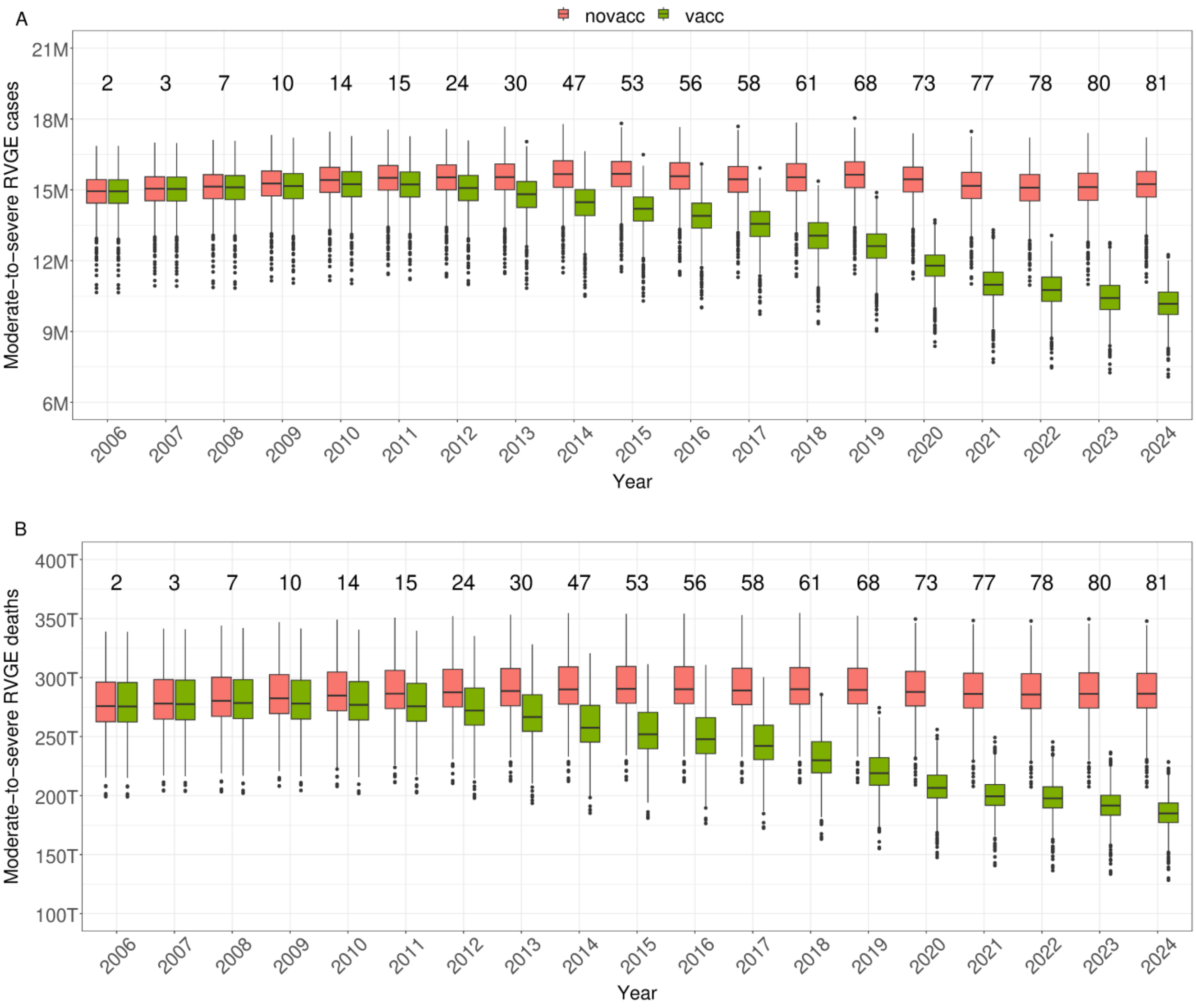


**S8 Fig. Annual boxplots of model-simulated moderate-to-severe RVGE cases (A) and RVGE-attributable deaths (B) across the 112 LMICs, with (green) and without (red) rotavirus vaccination, for the period 2006-2024.** Cases are presented in millions and deaths in thousands.

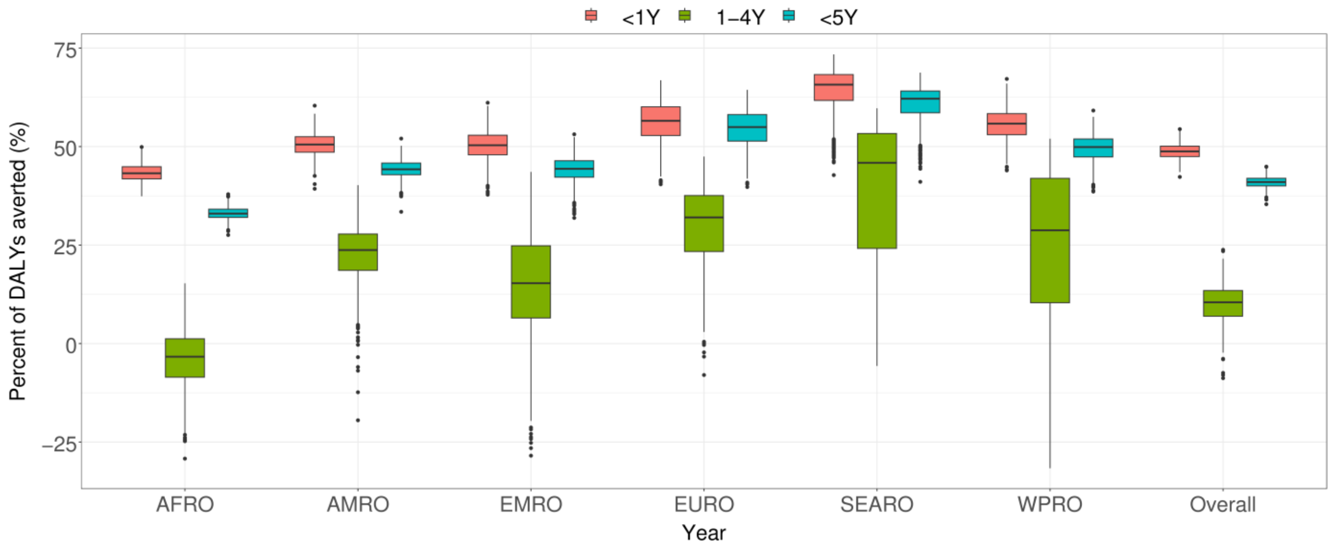


**S9 Fig. Percent of disability-adjusted life years (DALYs) averted by rotavirus vaccination, stratified by age group and WHO region, across the 81 countries that had introduced the vaccine by the end of 2024**. The overall represents the aggregated results for all 81 countries. Red, green, and blue colors represent age groups <1 year, 1-4 years, and <5 years, respectively. Estimates represent the proportional reduction in DALYs compared to a no-vaccination baseline.

**S3 Table. Estimated overall median and 95% uncertainty intervals of averted DALYs, stratified by age group, for historical and projected vaccination strategies (in millions).** For the historical estimates, the baseline is the no-vaccination simulations; for the projected strategies, the baseline is the current vaccination simulations; and for the additional dose scenario, the baseline is the current two-dose simulations.

| Strategies | <1 Year (95% UI) | 1-4 Years (95% UI) | <5 Years (95% UI) |
|---|---|---|---|
| **Historical (2006-2024)** | | | |
| Historical | 50 (42; 60) | 3 (0; 6) | 53 (45; 61) |
| **Projected (2025-2034)** | | | |
| Full adoption | 12 (9; 18) | 4 (3; 6) | 17 (13; 21) |
| Scaled-up coverage to 95% | 41 (33; 48) | 13 (10; 18) | 55 (45; 61) |
| Suspension | -62 (-79; -51) | -4 (-8; 1) | -66 (-80; -56) |
| **Third dose (2025-2034)** | | | |
| Third dose at current coverage | 7 (5; 10) | 4 (3; 6) | 11 (8; 15) |
| Third dose at 95% coverage | 16 (13; 20) | 8 (6; 10) | 24 (20; 28) |

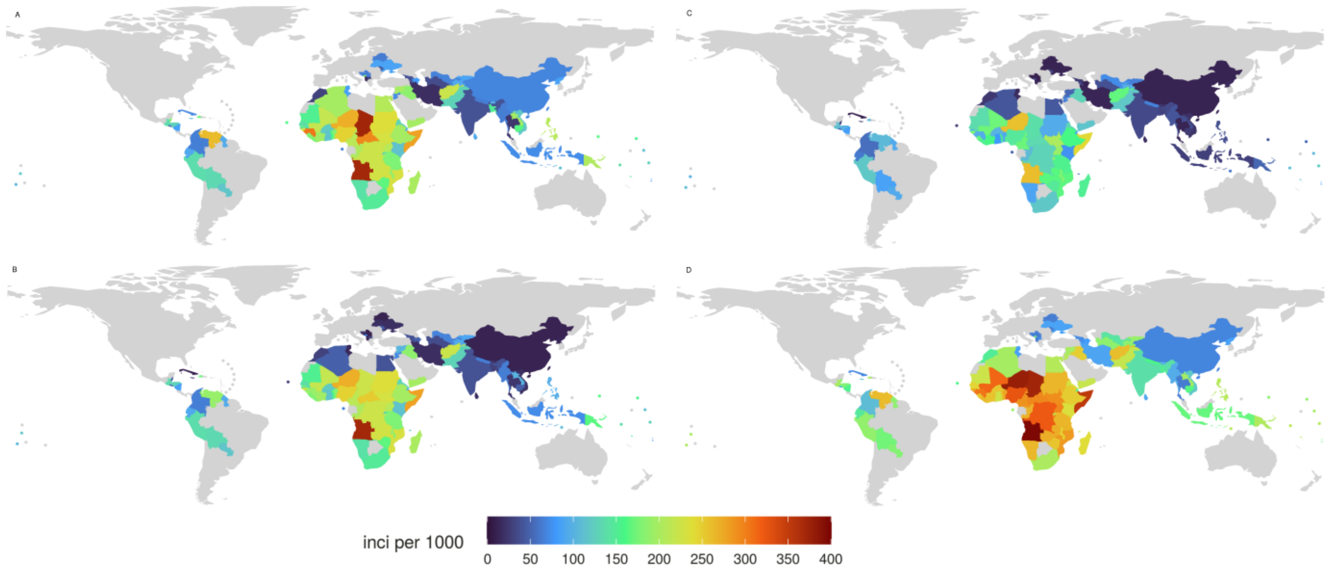


**S10 Fig. Model-projected median incidence of RVGE per 1,000 children under five years of age under different vaccination scenarios from 2025 to 2034**. (A) Current vaccination programs in 83 countries, (B) Current programs plus the introduction of a three-dose schedule with DTP3-based coverage in countries without vaccination, (C) same as (B), but with coverage scaled up to 95% in countries with low vaccination rates, and (D) No vaccination (suspension or no introduction in countries without a program).

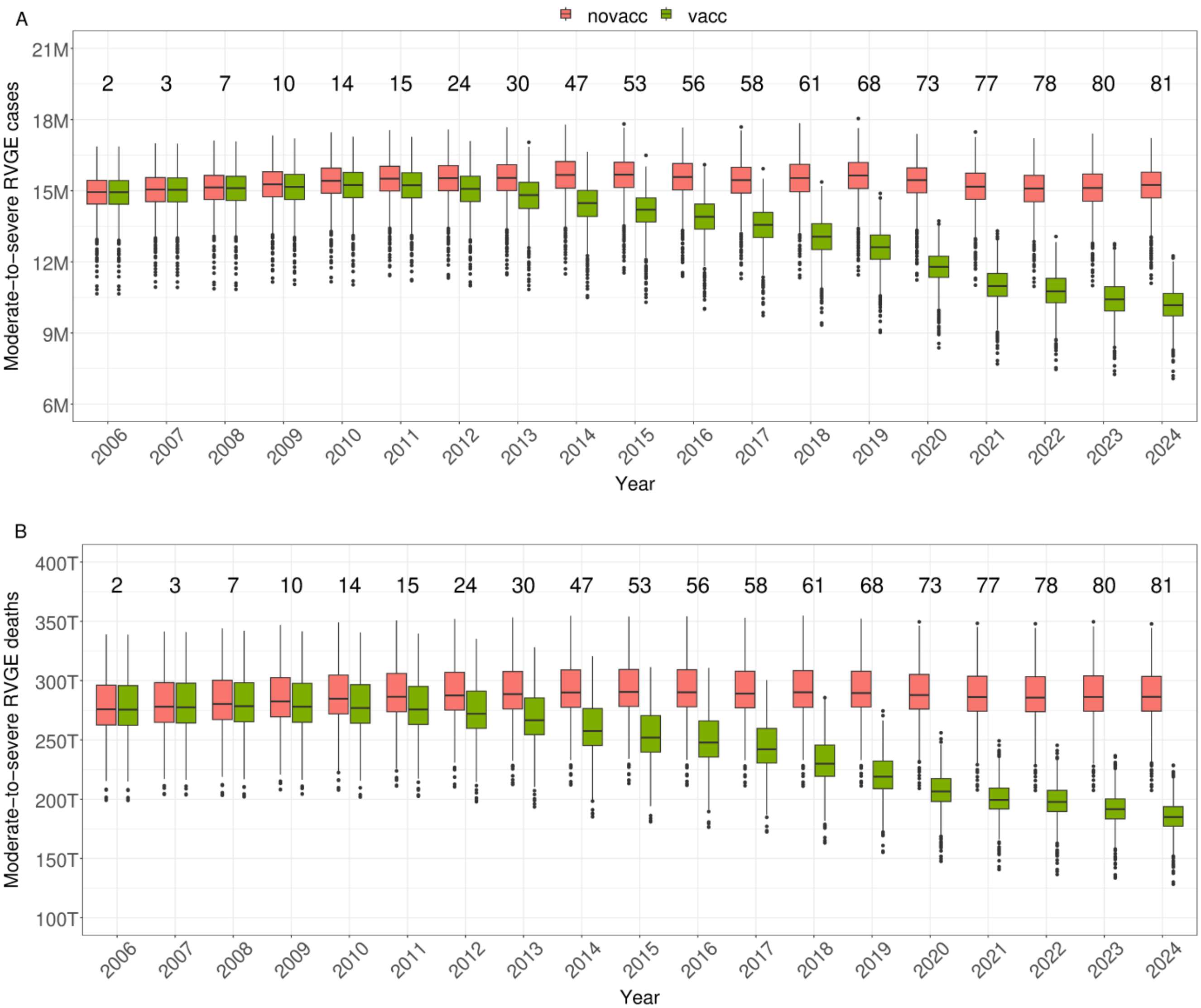


**S11 Fig. Annual boxplots of model-simulated moderate-to-severe RVGE cases (A) and RVGE-attributable deaths (B) across 112 LMICs for the period 2025–2034, under four different vaccination scenarios**. Cases are presented in millions and deaths in thousands. The red, green, blue, and purple colors represent the no-vaccination (suspension), current, full adoption, and 95% scaled-up coverage scenarios, respectively.

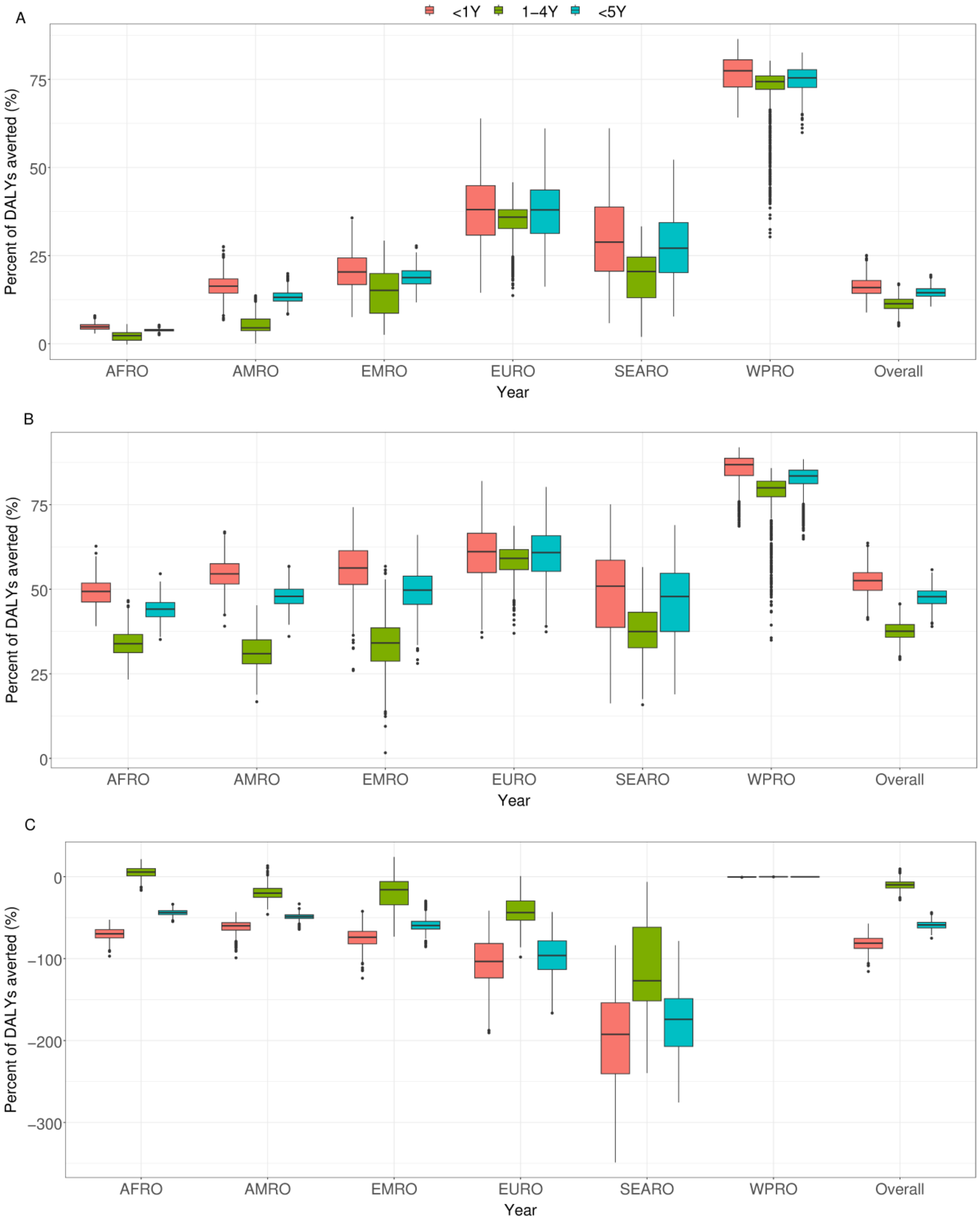


**S12 Fig. Percent of DALYs averted by different rotavirus vaccination scenarios, stratified by age group and WHO region, between 2025 and 2034**. The baseline for comparison is the current vaccination scenario, with results shown for full adoption (A), 95% scaled-up coverage (B), and suspension (C) for the 112 LMICs. Red, green, and blue colors represent age groups <1 year, 1-4 years, and <5 years, respectively.

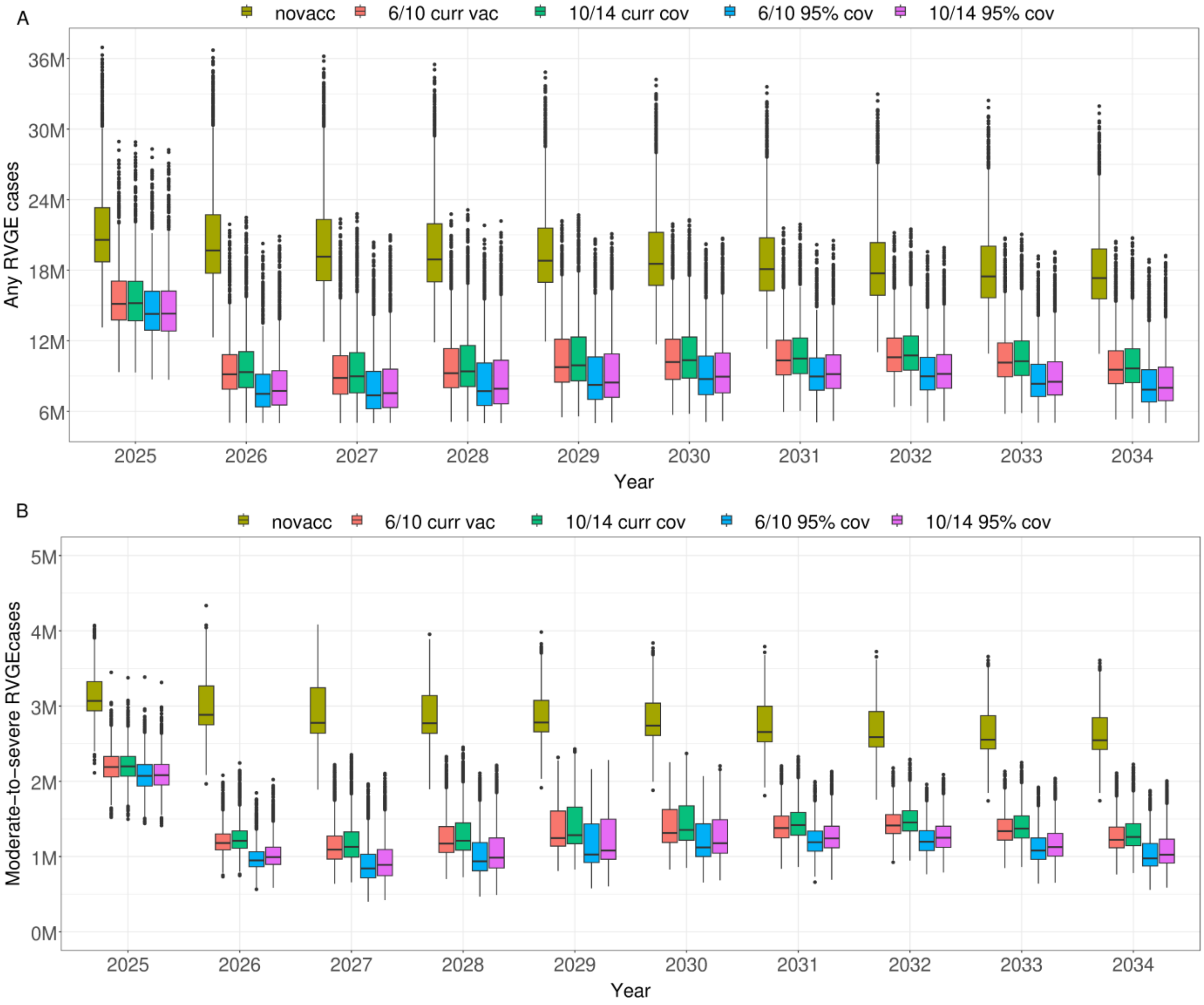


**S13 Fig. Projected impact of rotavirus vaccination on any (A) and moderate-to-severe (B) RVGE cases among children under 5 years of age across 29 LMICs over a 10-year period (2025-2034), in the absence of vaccination**. Results are presented for two commonly used two-dose schedules (6/10 weeks and 10/14 weeks) at both current DTP3 coverage and 95% scaled-up coverage.

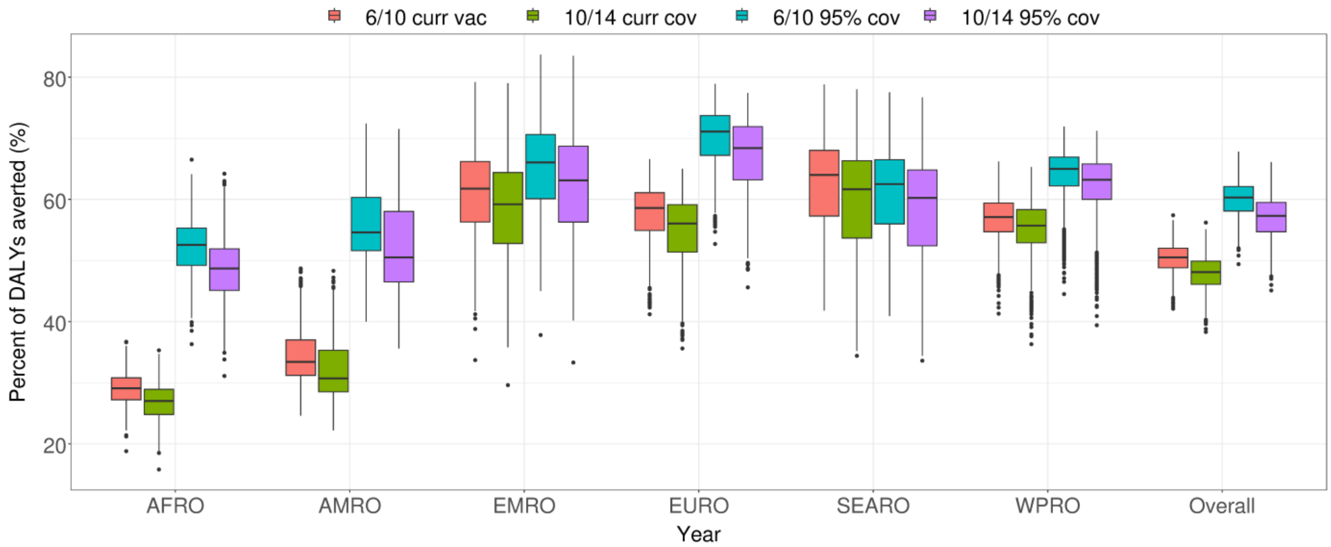


**S14 Fig. Percent of DALYs averted by introducing two-dose schedules (6/10- and 10/14-week) in 29 countries without vaccination, stratified by age group and WHO region, between 2025 and 2034**. The baseline for comparison is the no-vaccination simulations.

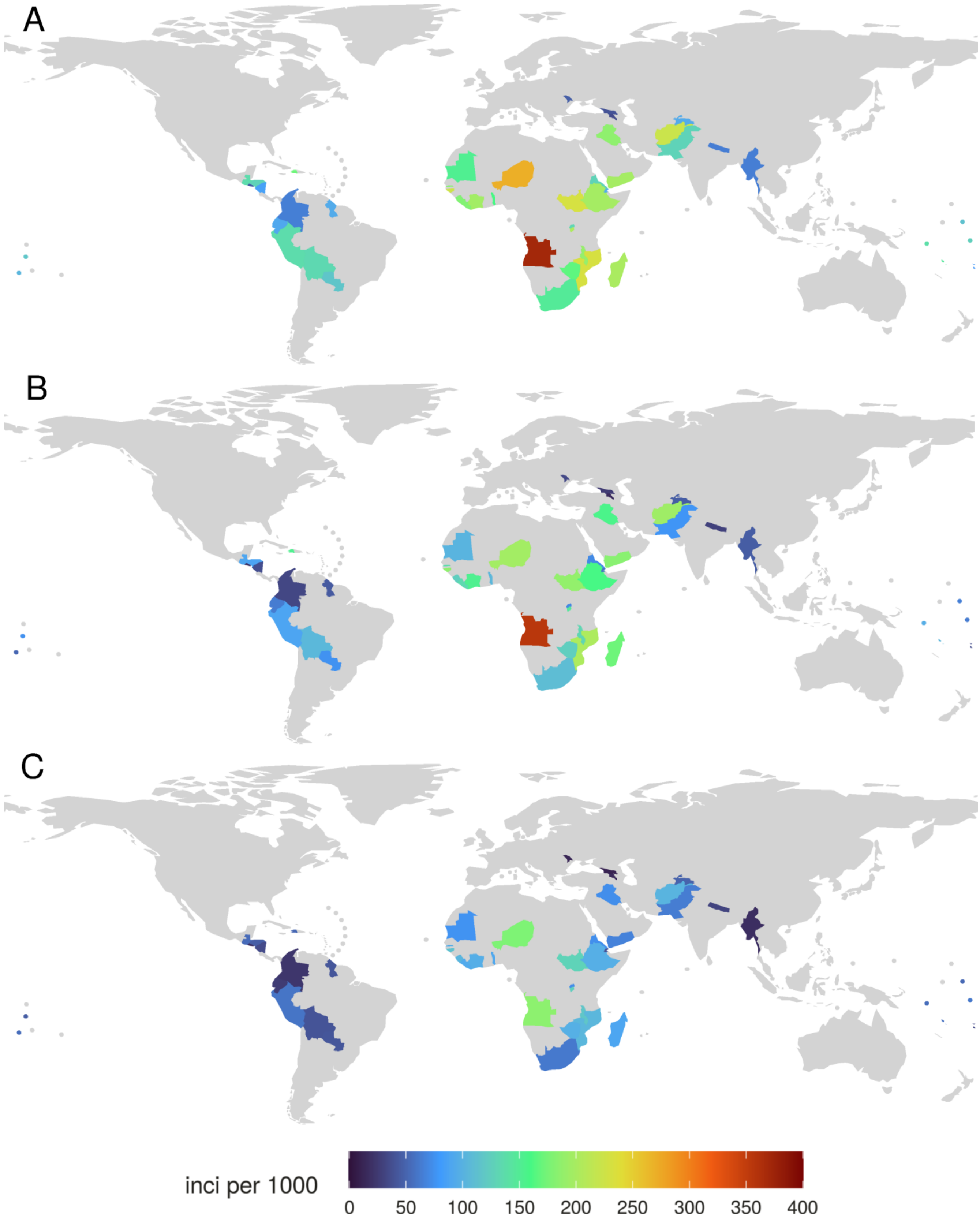


**S15 Fig. Model-projected median incidence of RVGE per 1,000 children <5 years of age when a third dose is added in countries currently using two-dose schedules**. (A) Current two-dose schedules, (B) Addition of a third dose at current coverage levels, and (C) Addition of a third dose with coverage scaled up to 95%.

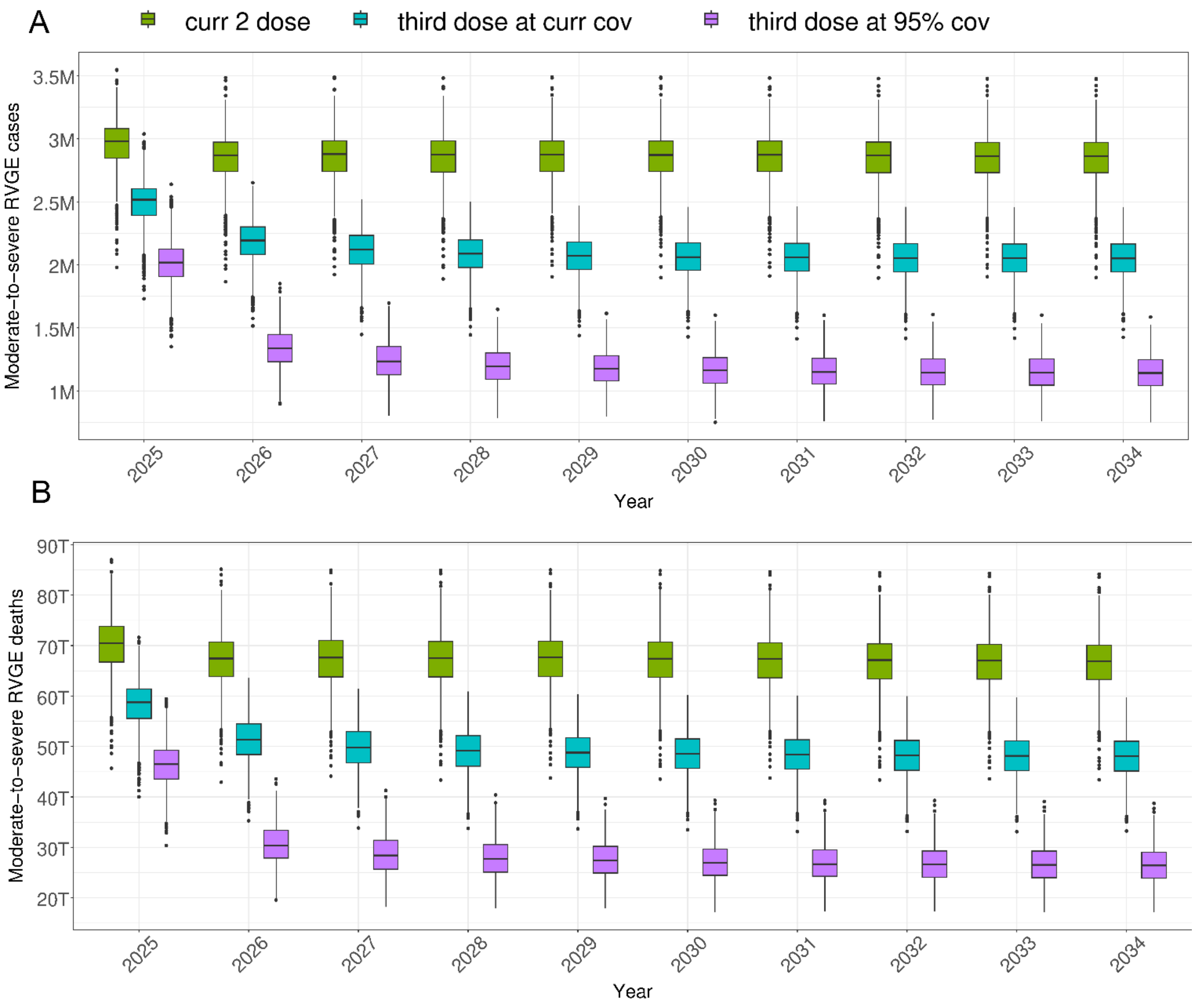


**S16 Fig. Annual boxplots of model-simulated moderate-to-severe RVGE cases (A) and RVGE-attributable deaths (B) across 112 LMICs for the period 2025–2034, under scenarios involving the addition of a third dose**. Cases are presented in millions and deaths in thousands. The green, blue, and pink colors represent the current two-dose scenario, the addition of a third dose at current coverage, and the addition of a third dose at 95% scaled-up coverage, respectively.

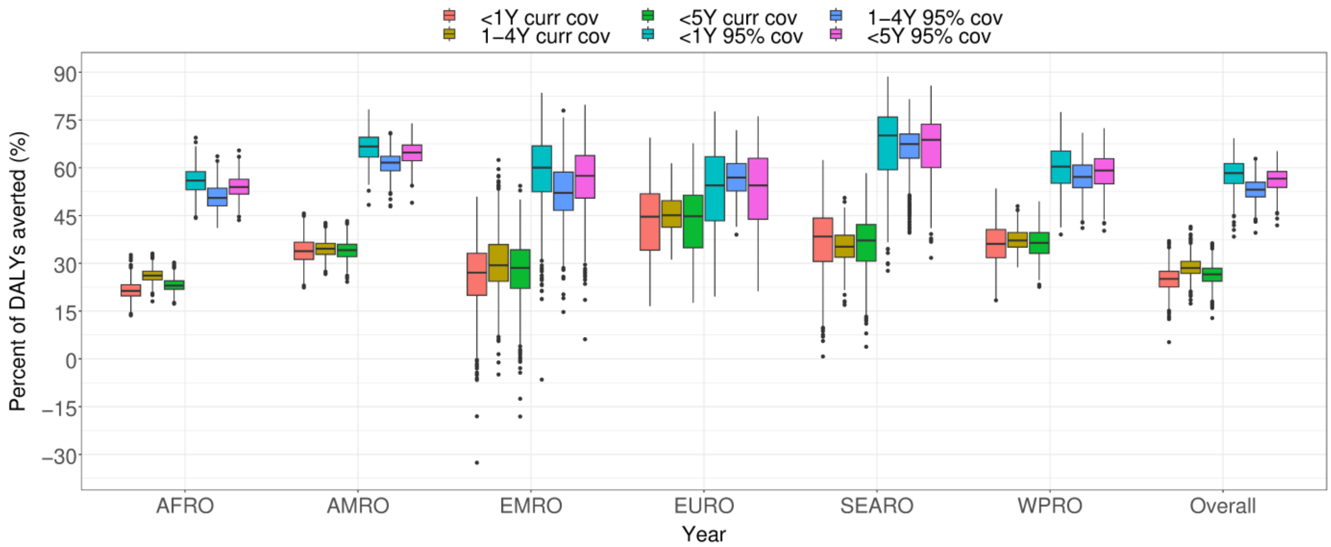


**S17 Fig. Percent of DALYs averted by implementing two-dose vaccination schedules, stratified by age group and WHO region, between 2025 and 2034**. The baseline for comparison is no-vaccination simulations. Results are shown for the introduction of 6/10-week and 10/14-week vaccination schedules, at both current and 95% scaled-up coverage, across 29 LMICs without vaccination.
